# Two weeks of cumulative tendon load monitored by insole sensors is associated with plantar flexor function in Achilles tendinopathy

**DOI:** 10.1101/2025.07.01.25330442

**Authors:** Ke Song, Michelle P. Kwon, Andy K. Smith, Ryan T. Pohlig, Karin Grävare Silbernagel, Josh R. Baxter

**Author notes:** Corresponding Author: Ke Song, PhD, Department of Orthopaedic Surgery, University of Pennsylvania 3737 Market Street, 6th Floor, Philadelphia, PA, USA 19104.

## Abstract

Tendon loading dictates rehabilitation outcomes in Achilles tendinopathy but is difficult to track in the real world. In this study, we used instrumented insole sensors to monitor Achilles tendon load for two weeks in fifteen individuals with Achilles tendinopathy, who also completed assessments of their plantar flexor strength, dynamic function, and survey-based outcomes. We used insole data to estimate two types of cumulative Achilles tendon load: overall (≥0.3×body weight) and high-level load (≥3×body weight). We determined Pearson correlations between (1) overall and high-level tendon loads, (2) plantar flexor moment, power, work, (3) heel raise height, repetitions, countermovement jump height, and (4) self-reported symptoms and activity. Overall cumulative tendon load moderately correlated to isometric plantar flexor moment (r = 0.543) and weakly to isokinetic and dynamic functions (0.128–0.413). Cumulative high-level tendon load strongly correlated to heel raise height (0.687) and fast isokinetic moment (0.625), and moderately to other functional measures (0.470–0.592). Symptoms weakly correlated to overall (0.392) and moderately to high-level load (0.436). Self-reported activity weakly correlated to overall (0.297) and strongly to high-level load (0.617). Stronger associations with the high-level Achilles tendon load than the overall load suggest that clinical function assessments provide insight into the real-world performance of high-loading activities. In contrast, the disconnect between overall tendon loading and plantar flexor function may explain the variability in recovery outcomes. Self-reported activity and standard heel raises represent high-level tendon load well, yet they do not always suggest functional deficit. Sensor-monitored tendon load shows promise as a new biomarker for real-world plantar flexor function in Achilles tendinopathy.

## Introduction

Achilles tendinopathy is a painful and debilitating chronic condition that often impacts physically active individuals [1,2]. Exercise progression to incrementally load the Achilles tendon is the clinical standard for rehabilitation and is effective in alleviating symptoms [3–7]. However, most rehabilitation programs are designed to progress Achilles tendon load through a prescribed exercise protocol but focus less on modifying the load outside of the exercise sessions, despite the evidence that prolonged underloading and overloading are both detrimental to tendon health [8,9]. This may explain the greatly varied treatment outcomes as 35–60% experience persistent pain and up to 50% seek alternative treatments, including surgery [10–12]. Some rehabilitation programs have thought to adjust tendon load incurred during daily living based on patient-reported pain and activity level [3,5]. Yet, self-reported activities are subjective and may not fully capture the actual status of real-world tendon loading. An unmet clinical need for improving treatment efficacy is to monitor Achilles tendon load during daily living and determine how it relates to symptom severity, self-reported activity, and plantar flexor function. Especially, continuous data that track real-world Achilles tendon loading status will provide mechanistic insights into how patient symptoms and function change overtime, which can improve clinician’s ability to customize exercise progression and daily activity modification. This requires quantifying the cumulative effect of Achilles tendon load resulting from all exercises and routine activities, and their associations with clinical outcome measures. However, quantifying the real-world Achilles tendon load over a long duration remains a major technical challenge, which impedes precision rehabilitation and warrants innovation.

Wearable sensing is a rapidly evolving novel approach for quantifying real-world physical activities and biomechanics. Sensors that measure body segment accelerations, temperature, heart rate, and other biomarkers are increasingly common [13–15]. Recent studies have explored using wearable motion sensors for physical activity monitoring in individuals with Achilles tendinopathy [16,17]. However, no sensor-based studies on this population have directly quantified mechanical load as a monitoring biomarker. Our group is using multiple wearable sensor paradigms to monitor Achilles tendon load in the real world [18,19], including instrumented insoles that directly measure and track plantar contact forces [19–22]. The purpose of our current study was to use insole sensors to monitor cumulative tendon load in individuals with Achilles tendinopathy and determine how cumulative loading is associated with plantar flexor function, symptoms, and self-reported activity. We hypothesized that insole sensor-measured Achilles tendon load would be strongly correlated to plantar flexor function and self-reported activity, while moderately or weakly correlated to symptom severity.

## Results

### Study participant characteristics and survey measures

Among our 19 initially recruited participants, fifteen (sex: 7 males, 8 females) successfully completed their two-week insole monitoring experiment (**Table 1**). These 15 participants reported highly variable Achilles tendinopathy severity, as their VISA-A score on the (more) symptomatic side ranged from 11 to 96 out of a 100-point scale. Their self-reported activity level was also highly variable, with all values (1–6) on the Physical Activity Scale observed.

**Table 1.** Study participant characteristics (n = 15, sex: 7 males, 8 females) and self-reported survey measures.

*Study participant characteristics and survey measures*
|  |  |  |  |  |  |
| --- | --- | --- | --- | --- | --- |
| **Age**<br>(years old) | **Height**<br>(m) | **Mass**<br>(kg) | **BMI**<br>(kg/m<sup>2</sup>) | **PAS**<br>(1 – 6 scale) | **VISA-A**<br>(0 – 100 scale) |
| 46.7 ± 12.0<br>(34 – 70) | 1.72 ± 0.12<br>(1.55 – 1.91) | 95.6 ± 24.9<br>(59.0 – 140.6) | 32.9 ± 9.9<br>(20.6 – 54.8) | 3.8 ± 1.6<br>(1 – 6) | 54.2 ± 26.1<br>(11 – 96) |
| Data presented in mean ± 1 standard deviation (minimum – maximum). BMI, body-mass index; PAS, Physical Activity Scale; VISA-A, Victorian Institute of Sports Assessment – Achilles questionnaire. |  |  |  |  |  |

### Cumulative Achilles tendon load: descriptive results from insole sensors

Study participants used the instrumented insole on an average of 10.1 days (standard deviation, SD: 2.2 days, range: 6–14) during their two-week monitoring period. On average, participants spent time loading their Achilles tendon above the overall level for a total of 24.1 hours (SD: 14.3 hours, range: 10.1–61.6) and accumulated 24.2×BW×hour of overall Achilles tendon load (SD: 15.7×BW×hour, range: 8.1–62.1). As a subset of this overall load, participants spent time loading their Achilles tendon above the high-level threshold for a total of 0.6 hours (SD: 0.7 hours, range: 0–2.4) and accumulated 2.3×BW×hour of high-level tendon load (SD: 2.8×BW×hour, range: 0–9.3). These *non-normalized* cumulative hours and loading impulses are dependent on the total insole wear time by each participant. When divided by the total loading time with the insole (i.e., hours spent above 0.3×BW), participants accumulated 0.98×BW of *normalized* Achilles tendon load *per hour* above the overall level (SD: 0.18×BW, range: 0.69– 1.27) and 0.08×BW *per hour* beyond the high level (SD: 0.07×BW, range: 0–0.21).

### Cumulative tendon load vs. plantar flexor functional capacity (dynamometer)

Dynamometer-based measures of plantar flexor functional capacity generally had moderate correlations with the overall cumulative Achilles tendon load, and moderate-to-strong correlations with the high-level tendon load, depending on the testing condition (**Fig. 1**, **Supplementary Fig. S1**). Specifically, the overall tendon load *per hour* was weak-to-moderately and positively correlated to all plantar flexor moment, power, and mechanical work (Pearson r = 0.327–0.543; **Fig. 1A**). The high-level tendon load *per hour* had moderate (and relatively stronger) positive correlations to most dynamometer-based plantar flexor functional capacity (r = 0.475– 0.592; **Fig. 1B**). The peak plantar flexor moment during fast isokinetic contraction met our a priori criteria for a strong correlation (r = 0.625; **Fig. 1B**, top right).

**Figure 1.**
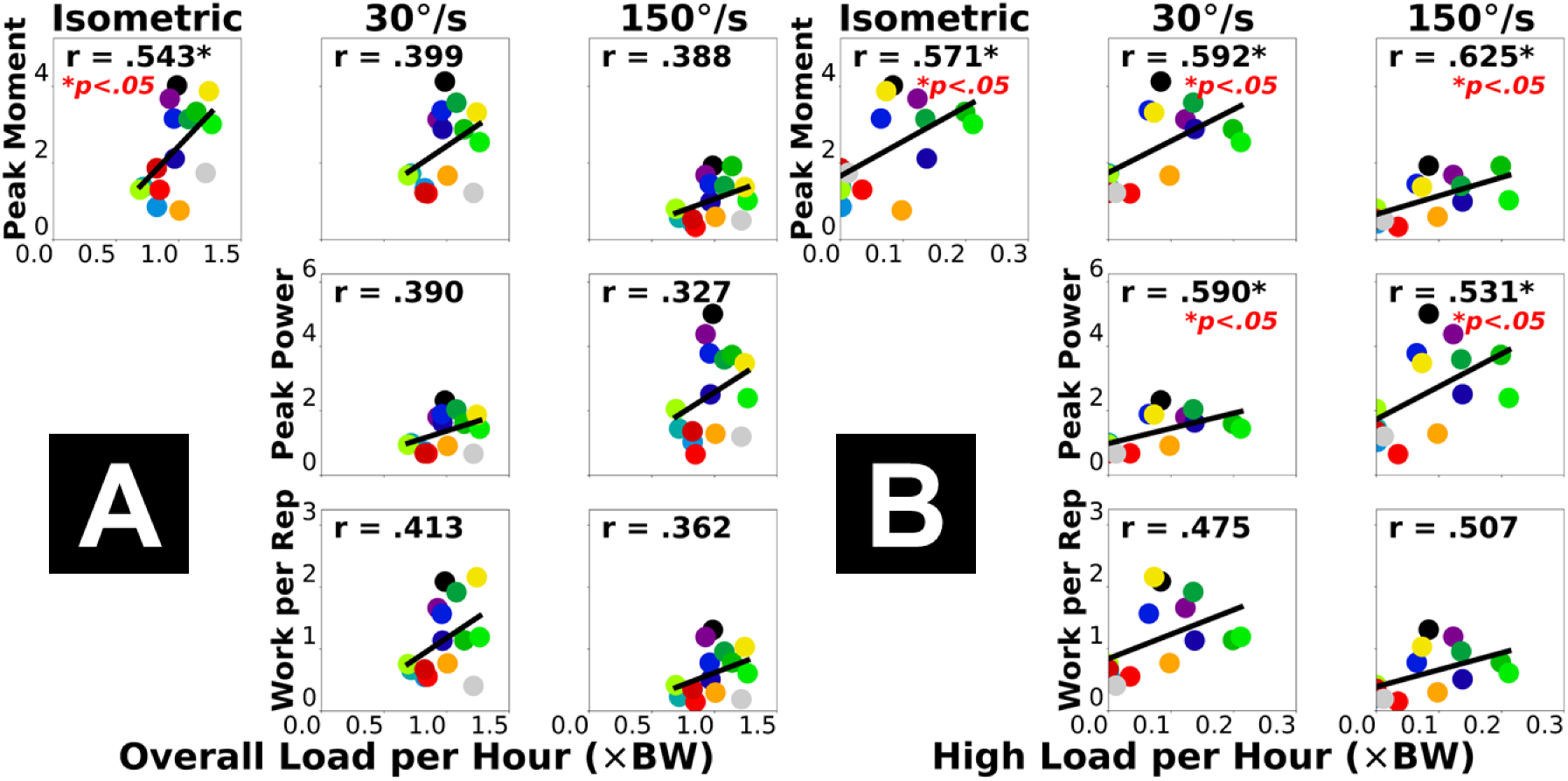
Pearson correlations between dynamometer-based plantar flexor functional capacity (vertical axis) and insole-based cumulative Achilles tendon load (horizontal axis): normalized overall load *per hour* **(A)** and high-level load *per hour* **(B)**. Each dot represents a participant. “*” annotates statistical significance of the correlation (*p* < 0.05).

### Cumulative tendon load vs. plantar flexor dynamic function (motion capture)

Motion capture-based measures of plantar flexor dynamic function had weak correlations with the overall cumulative Achilles tendon load (**Fig. 2A**) and moderate-to-strong correlations with the high-level tendon load (**Fig. 2B**). Across the three exercises, double-leg heel raise height had the strongest correlations with both insole-based cumulative tendon load measures (**Fig. 2**, top row), with a strong correlation to the high-level tendon load *per hour* (r = 0.687).

**Figure 2.**
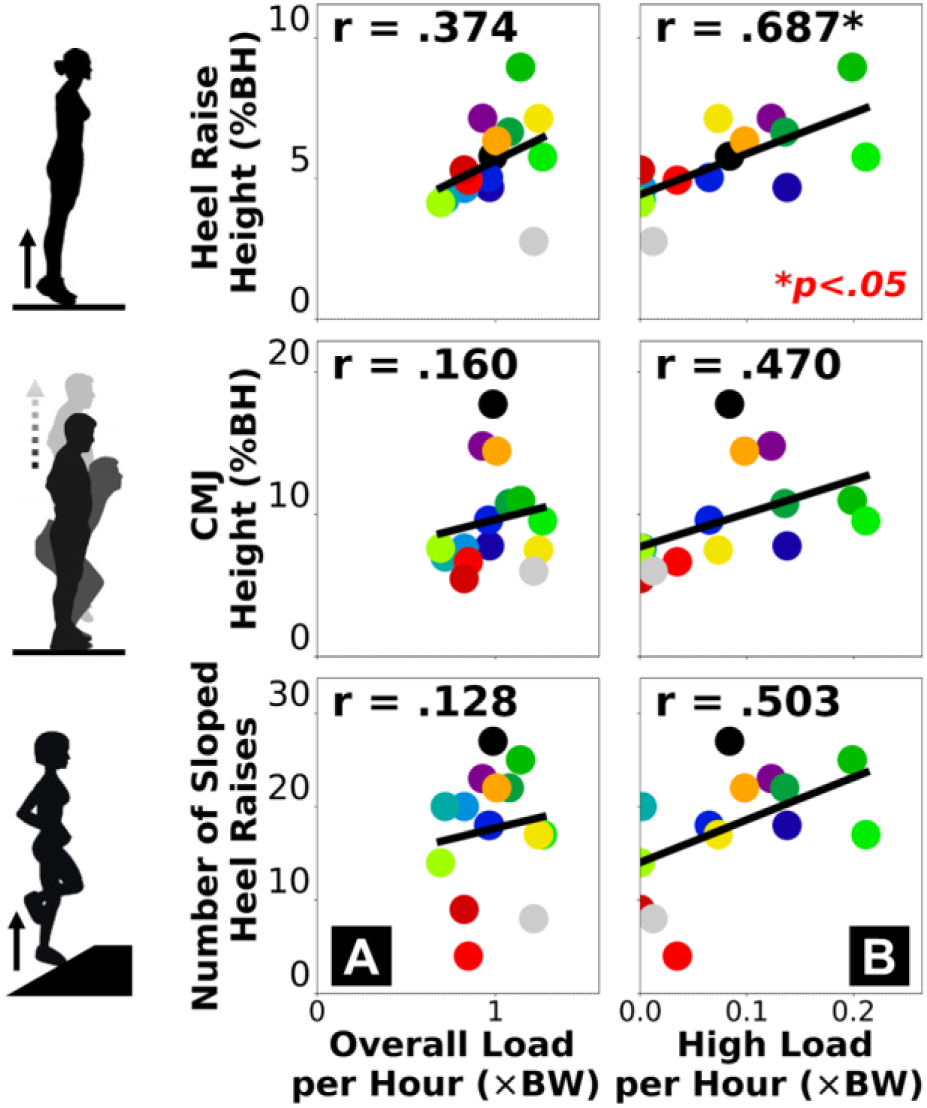
Pearson correlations between motion capture-based plantar flexor dynamic function (vertical axis) and insole-based cumulative Achilles tendon load (horizontal axis): normalized overall load *per hour* **(A)** and high-level load *per hour* **(B)**. Each dot represents a participant. “*” annotates statistical significance of the correlation (*p* < 0.05).

### Cumulative tendon load vs. self-reported clinical measures (surveys)

Overall cumulative Achilles tendon load *per hour* had weak positive correlations with all three self-reported survey measures: age (r = 0.204), severity of Achilles tendinopathy by the VISA-A score (r = 0.392), and current activity level by the PAS score (r = 0.297) (**Fig. 3A**). Cumulative high-level Achilles tendon load *per hour* had a weak negative correlation to participant age (r =-0.330), moderate positive correlation to the VISA-A score (r = 0.436), and strong positive correlation to the PAS score (r = 0.617) (**Fig. 3B**).

**Figure 3.**
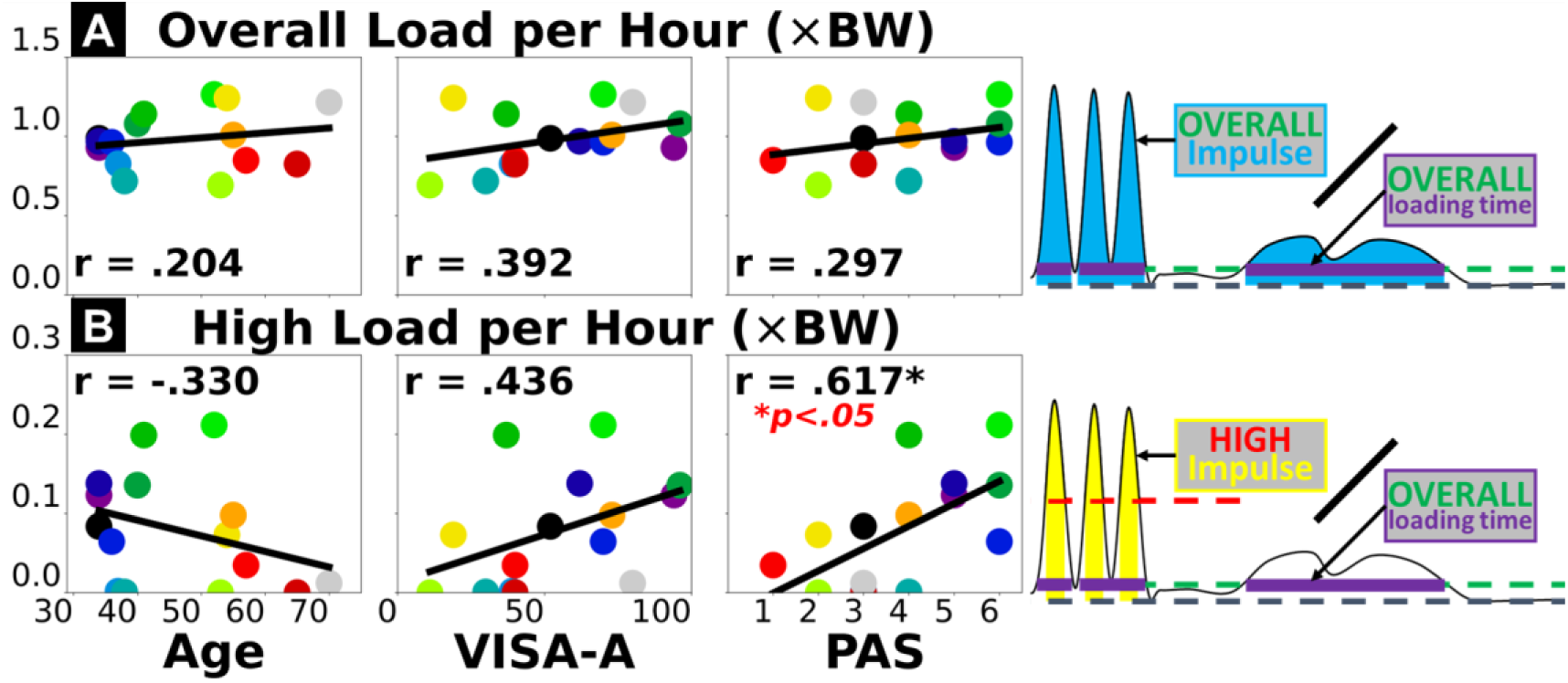
Pearson correlations between insole-based cumulative Achilles tendon load (vertical axis) and survey-based measures of participant characteristics or outcomes (horizontal axis). Insole-based measures include normalized overall load *per hour* **(A)** and high-level load *per hour* **(B)**. Survey-based measures include age **(left)**, severity of Achilles tendinopathy – VISA-A score **(center)**, and self-reported current activity level – PAS score **(right)**. Each dot represents a participant. “*” annotates statistical significance of the correlation (*p* < 0.05).

### Effects of insole data subsampling

Using reduced days of insole data led to increased error in estimating the overall cumulative and high-level tendon loads (**Fig. 4**, **Supplementary Fig. S2**). Estimates of the overall tendon load were relatively robust with subsampling, as the mean absolute percent error remained <10% even when only using 1 day of insole data (**Fig. 4A & 4C**). In contrast, using 1 day of data led to misestimated high-level load *per hour* for some participants on multiple days, resulting in a >60% mean absolute percent error across the group (**Fig. 4B & 4C**). This error was gradually reduced when increasing the insole sensor monitoring days. When using the first 6 days of data from each participant, mean absolute percent error of the high-level tendon load was 25.1% (**Fig. 4C**) with correlation r = 0.95 against the full dataset-based estimate (**Fig. 4D**).

**Figure 4.**
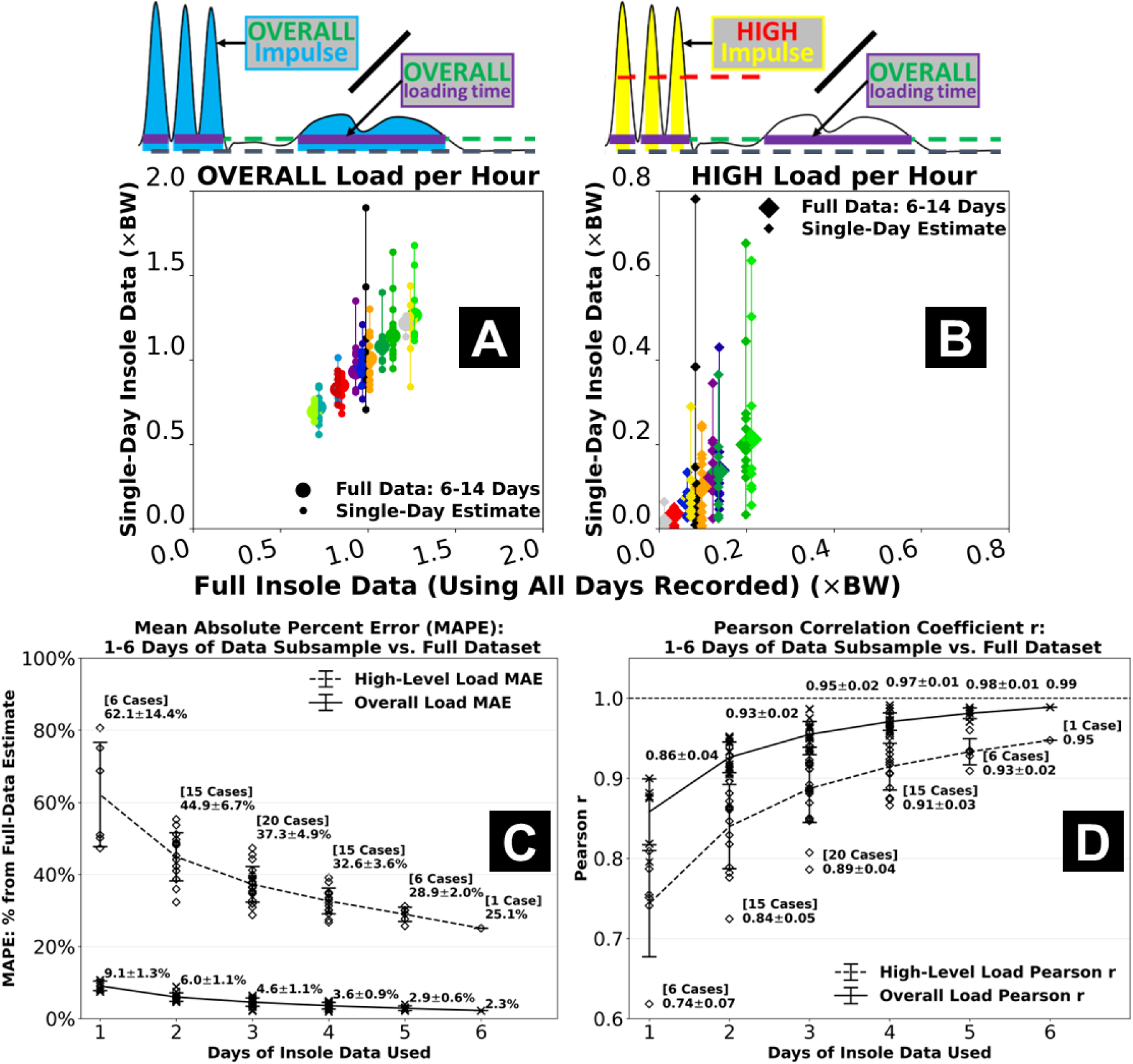
Effects of using a reduced number of days of insole data compared to using the full two-week dataset. Estimates from a single day of data often tended to under-or over-estimate the overall load **(A)** and especially the high-level Achilles tendon load *per hour* **(B)**. Mean absolute percent error for 1 day-based estimate was <10% from full dataset-based for the overall load, but >60% for the high-level load which also showed more day-to-day variations **(C)**. Using more days of data gradually reduced error and increased its correlations to the full dataset. When using the first 6 days of data, mean absolute percent error of the high-level load was 25.1% and correlation was r = 0.95 against the full dataset-based estimate **(D)**. Error bars represent ±1 standard deviation from the mean across all 1, 2,…, or 6 day-based estimates.

## Discussion

The purpose of this study was to use insole sensors to monitor cumulative tendon load in individuals with Achilles tendinopathy and determine how cumulative loading is associated with plantar flexor function, symptoms, and self-reported activity. We found that 6–14 days of Achilles tendon load monitored by the force-sensing insole, especially cumulative high tendon load beyond the level of walking, is moderate-to-strongly correlated to plantar flexor function. Consistent with our hypothesis, sensor-based tendon load estimates demonstrated stronger correlations to plantar flexor function yet weaker correlations to survey-based measures like symptoms and age. Cumulative high-level tendon load is strongly correlated to self-reported activity, which also supports our hypothesis. However, while self-reported activity is generally in agreement with objectively measured tendon load, they do not capture the same biomechanical information. Our new findings support the clinical value and advantage of using wearable sensors to monitor Achilles tendon load in daily living, for quantifying the real-world plantar flexor function in individuals diagnosed with Achilles tendinopathy.

Stronger associations with the high-level Achilles tendon load than the overall tendon load suggest that clinical function assessments for Achilles tendinopathy provide insight into the real-world performance of high loading activities. Cumulative high tendon load beyond the level of walking showed strong associations with both the functional capacity (**Fig. 1B**) and dynamic function (**Fig. 2B**) of the plantar flexors. These associations reached our criteria of strong (r ≥ 0.6) [23,24] for peak moment during fast isokinetic contraction (our most dynamic strength testing condition) and double-leg heel raise height. This indicates plantar flexor weakness for performing dynamic movements likely limits high-level tendon loading in the real world. Considering recent clinical findings that functional goals and recovery trajectories vary among patients with Achilles tendinopathy [25,26], for those who wish to return to sports or plyometric exercises, achieving a good level of cumulative high tendon load in daily living may be a useful clinical goal. Conversely, performing high loading activities regularly may protect patients against plantar flexor functional loss. Patients who have maintained a high tendon load level are unlikely to be impaired by weakness, and they may instead benefit from tendon load management through daily activity modifications for reducing symptoms. As opposed to the high-level load, the overall cumulative tendon load did not strongly associate with plantar flexor function (**Fig. 1A & 2A**). Isometric moment was the only functional measure moderately correlated to overall tendon load (r = 0.543), while all isokinetic and dynamic measures had weaker correlations (r ≤ 0.413). This means overall tendon loading in daily life at most partially reflects isometric muscle strength but cannot explain plantar flexor dynamic functionality, which indicates a potential disconnect between tendon health status and the real-world overall loading. For example, some (but not all) patients may expose their Achilles tendon to detrimental overloading or underloading in daily living that mismatches their functional recovery progress [8,9], which could contribute to suboptimal recovery and thus explain the greatly varied treatment outcomes among patients that clinicians observed [10–12].

Survey-based measures also had stronger associations with the high-level Achilles tendon load compared to the overall tendon load. The correlations between overall tendon load and age, symptoms, and self-reported activity were all weak (**Fig. 3A**), indicating that overall tendon load attributed to general daily living activities is neither fully explained by older age or a more sedentary lifestyle, nor adequately explains worse symptoms. A possible reason is that walking is the most common daily movement for most individuals, yet those who are of older age or self-perceive as physically inactive do not necessarily walk less. Age was negatively correlated to the high-level tendon load (r =-0.330, **Fig. 3B**) likely due to older individuals engaging in fewer dynamic activities, but this weak association is insufficient to confirm age as a reliable biomarker for reduced high-level Achilles tendon load either. Finally, weak-to-moderate associations with the VISA-A score indicate that symptoms like pain may not fully explain how Achilles tendon loading status changes in daily living (or vice versa). Confirming the cause-effect relationship between symptoms and tendon loading is beyond the scope of this cross-sectional study and warrants future longitudinal research.

Self-reported activity and standard heel raises both represented high-level tendon load well, yet they did not always suggest plantar flexor functional deficit. For our study participants, the cumulative high-level Achilles tendon load was strongly correlated to self-reported current activity level (**Fig. 3B**) and the double-leg heel raise height (**Fig. 2B**). Comparing across exercises, we found real-world tendon loading to be better associated with double-leg heel raise height compared to countermovement jump height and inclined single-leg heel raise repetitions (**Fig. 2**). This may be partly due to more variable techniques adapted by our heterogeneous group of study participants (**Table 1**) to perform more dynamic and demanding tasks like jumping. The double-leg heel raises are easier to perform and standardize, which may explain the consistent relationship to real-world tendon loading status in a heterogeneous patient population. Generally, our data supports the clinical usefulness of patient-reported activity surveys for informing exercise progression, and standard heel raises as a reliable rehabilitation exercise to improve planter flexor function [3–7]. However, they only marginally exceeded our criteria for strong correlation (r = 0.614–0.687) and do not fully explain real-world tendon loading. Despite being easier to implement than wearable sensors, a shortcoming of survey-based activity monitoring is that the discrete response options may be interpreted differently across individuals, inducing subjectivity and impeding specificity. For example, 3 of our participants responded the highest activity level (PAS = 6), but their cumulative high-level tendon load was either below, at, or above the group-wise trend in a divergent manner (**Fig. 3B**, bottom right). In a supplementary analysis, we found that the plantar flexor functional measures had weaker correlations to the PAS score (r = 0.227–0.491, **Supplementary Fig. S3 & S4**) than to insole-based high-level load (r = 0.470–0.687, **Fig. 1B & 2B**). This finding indicates that self-reported activity in Achilles tendinopathy patients may mismatch whether they have fully regained plantar flexor function. Instead, sensor-monitored cumulative Achilles tendon load better represents the capacity and dynamic function of the plantar flexors, therefore shows promise as a useful new biomarker for real-world plantar flexor function in physically active individuals.

To our knowledge, this study is the first to monitor foot contact force and estimate Achilles tendon load over a weeks-long duration. We chose a two-week paradigm to capture as much real-world insole data as we could and explore their “best scenario” associations with clinical measures. This allowed us to evaluate whether insole-based tendon load monitoring is clinically meaningful and worthy of future development. Although the associations with plantar flexor function suggest insole-based measures have promising clinical value, securing the multi-day insole data had been a challenging process for both our study participants and our research team. Practical constraints included but were not limited to shoe type, preference about wearing shoes at home, and concerns about the insole sensor appearance. These constraints all limited our participants’ ability to wear the insole and contributed to the varied actual days (6–14) and hours each day they logged data. In our subsampling analysis, we found that when only using 1 day of insole data, error against the full dataset remained low for the overall tendon load estimate (<10%) but was higher for the high-level load (>60%) (**Fig. 4C**). One day of insole sensor data may thus be sufficient to reliably characterize habitual tendon loading required for daily living mobility. Instead, multi-day insole monitoring may be necessary to ascertain the high-level tendon loading status, including its day-to-day variations, that are unique to sports and dynamic activity. We expect the challenges we experienced to impact most studies that implement day(s)-long insole sensor monitoring. We thus recommend researchers to carefully choose insole monitoring durations based on the population studied and standardize monitoring protocols. These considerations will help researchers expand cohort size and discover clinically relevant loading parameters with a higher level of confidence.

Beyond monitoring duration, researchers should also be aware of several other technical and practical challenges before using instrumented insoles for real-world monitoring in scale. For example, technical factors like data storage capacity and the need to rechange sensors may limit the monitoring duration and add to data inconsistency. Another standing challenge is the technical drawbacks of the instrumented insoles, such as signal noise, nonphysical artifacts, and device malfunctions vulnerable to insole misfit or miscalibration (**Supplementary Fig. S6 & S7**). Future development work should mitigate these limitations by improving sensor storage capacity, battery life, and technical robustness. Combined with wear time variability, we had to assume that the total loading time with the insole that we captured were representative of wearers’ typical activity and Achilles tendon loading status. If a study participant would only wear the insole whenever they exercise, we would overestimate their cumulative tendon load *per hour* on those days (**Fig. 4**). Conversely, if the insole monitoring day for a runner coincided with their resting day of the week, we would underestimate their habitual high-level tendon load. We urge our fellow researchers to continue exploring better strategies to reduce participant burden and improve study protocol adherence, for capturing more consistent and more representative insole data. Researchers should also explore the utility of other wearable sensors that have established good feasibility for continuous monitoring, including inertial measurement units [13,16–18], and identify how sensor-based measures of motion (body acceleration, velocity, position, orientation, etc.) associate with relevant mechanical biomarkers (e.g., Achilles tendon load) and clinical outcome measures.

Our findings should be considered with several important limitations. First, while dividing by the total loading time partly addressed the variability in wear time and study protocol adherence, our *normalized* measures of cumulative loading may be difficult to contextualize or implement as a clinical measure. We elaborated our considerations and recommendations for future work in the previous paragraphs. Second, because the 0.3×BW baseline cutoff removed all transient unloaded periods including the swing phase of walking and running, our definition of the total loading time does not readily represent full active time of the insole wearer. Past studies have developed epoch-based algorithms to determine total active time wearing motion sensors [27], which can be adapted for evaluating insole sensor wear time and adherence to the monitoring protocol. Third, we did not record pain during or after the in-lab functional assessments. In the recent clinician consensus [28], two of the core outcome measures for Achilles tendinopathy are pain during and after functional activities, which should be included in future studies. Fourth, the interaction between plantar flexor function and tendon structure in Achilles tendinopathy is clinically recognized [3,25,26], but the relationships between plantar flexor muscle-tendon structure and cumulative Achilles tendon load remain unknown. Our ultrasound-based studies to investigate this question are ongoing. Finally, our sample size is small partly due to the technical and practical challenges with insole monitoring. Recent clinical studies have identified homogeneous subgroups within Achilles tendinopathy patients who likely need distinctive intervention strategies [25,26]. The stronger associations we observed in high-level tendon load indicate that cumulative Achilles tendon load may be of higher importance to the activity-dominant patient subgroup [25,26], but this requires confirmation in a larger cohort. The sample size of this study is insufficient to compare real-world Achilles tendon load among patient subgroups or stratify them according to our new insole-based measures. Our larger study is enrolling more patients for insole monitoring to address this aspect. Nonetheless, our current study takes a groundbreaking step by providing a first-of-kind biomechanical dataset that confirmed the clinical value and advantage of real-world wearable sensor monitoring.

In conclusion, cumulative Achilles tendon load monitored by instrumented insole through 6–14 days is moderate-to-strongly associated with plantar flexor function in individuals with Achilles tendinopathy. Stronger associations with the high-level Achilles tendon load than the overall load suggest that clinical function assessments for Achilles tendinopathy provide insight into the real-world performance of high loading activities. In contrast, the disconnect between overall tendon loading and plantar flexor function may explain the variability in recovery outcomes. Self-reported activity and standard heel raises represent high-level tendon load well, yet they do not always suggest plantar flexor functional deficit. Sensor-monitored Achilles tendon load shows promise as a useful new biomarker for real-world plantar flexor function in physically active individuals. Considering the associations between cumulative tendon loads and clinical measures, yet the standing challenges with using insole sensors, we advocate for further research and development to improve the practicality of insole monitoring paradigms and upscale analyses on real-world data. This will lead to a better understanding on the roles of tendon loading in real-world functional performance in individuals with Achilles tendinopathy.

## Methods

### Study participants

We recruited 19 adults with clinically confirmed Achilles tendinopathy from the University of Pennsylvania orthopaedic clinic and our local community. Our eligibility criteria were between 18 and 70 years of age, no prior Achilles tendon rupture, no other musculoskeletal injuries in the past six months, no use of nicotine or steroid medication, and not pregnant. A board-certified physical therapist (A.K.S.) confirmed the diagnosis of either midportion or insertional Achilles tendinopathy using patient history, palpation, and the Arc sign and Royal London Hospital tests [29]. Our study was approved by the Institutional Review Board at the University of Pennsylvania (IRB Protocol #850424) and all participants provided written informed consent before starting the experiments. We performed all research activities in this study in accordance with the Declaration of Helsinki and relevant human participant research regulations and guidelines.

### In-lab experiments: self-reported outcomes and plantar flexor function

Participants completed digital questionnaires in the REDCap database system (version 13, REDCap Project, Nashville, TN) [30,31]. These questionnaires surveyed participant age, sex, body height and mass, and the symptomatic side (more symptomatic side if bilateral). Participants reported the severity of their Achilles tendinopathy specific to the (more) symptomatic tendon using the 8-item Victorian Institute of Sports Assessment – Achilles (VISA-A) questionnaire [32], which is scored 0–100 with 100 indicating no symptom or disability. Participants also reported their perceived current physical activity level using the clinically validated Physical Activity Scale (PAS) [3,33], a single question scored 1–6 with 6 indicating regular intensive exercises several times a week and 1 indicating hardly any physical activity.

We assessed two types of plantar flexor function: capacity and dynamic function. We first quantified functional capacity of the plantar flexors via isometric and isokinetic strength testing. Participants were positioned prone on a treatment plinth adjacent to an isokinetic dynamometer (VAC System 4 Pro, Biodex Medical Systems, Shirley, NY) following our published experimental protocol [34]. Each participant performed three maximal voluntary contraction trials on their (more) symptomatic side, with three repetitions each: (1) isometric at neutral ankle position, (2) slow isokinetic (30°/s), and (3) fast isokinetic (150°/s). We quantified 7 measures of plantar flexor functional capacity, each normalized by body height times weight: peak moment during all 3 contraction types (unit: %H×W); peak power during slow and fast isokinetic contractions, by moment times plantarflexion speed (unit: %H×W×rad/s); mechanical work per slow and fast isokinetic contraction, by the integral of power over time (unit: %H×W×rad). We averaged each functional capacity measure across the 3 contraction repetitions.

We next quantified plantar flexor dynamic function using motion capture. Each participant performed three exercises commonly used in Achilles tendinopathy clinical assessment or treatment [3–7,35] in the following order: 10 double-leg heel raises, 5 single-leg countermovement jumps, then consecutive single-leg heel raises on a 25° inclined slope until 30 repetitions or physical exhaustion. We recorded videos of each exercise using 8 synchronized high-resolution cameras (2.1-megapixel Optitrack Prime Color, NaturalPoint Inc., Corvallis, OR), from which we quantified the shank and foot positions using a markerless motion tracking algorithm (Theia3D v2023.1.0.3160, Theia Markerless Inc., Kingston, ON, Canada) [36,37]. We calculated three clinically relevant kinematic measures of the plantar flexor dynamic function on the (more) symptomatic side [3–7,35]: double-leg heel raise height, single-leg countermovement jump height, and total repetitions of single-leg heel raises on the 25° slope. We normalized heel raise height and countermovement jump height by body height (unit: %BH) to account for participant body size differences [38–40], then averaged them across movement repetitions.

### At-home experiment: instrumented insole monitoring of Achilles tendon load

We used an instrumented force-sensing insole (Loadsol Pro 2 HMF, Novel GmbH, Munich, Germany) to monitor Achilles tendon load for up to two weeks. Each insole has 3 plantar force sensor pads (heel, midfoot, forefoot) embedded within its flexible surface. They are wire-connected to a miniature data logger that can be mounted on shoelaces and continuously log plantar contact forces with a battery life of ∼25 hours (**Fig. 5A**). These instrumented insoles use a smartphone application to begin and stop data acquisition and store them on the data logger.

**Figure 5.**
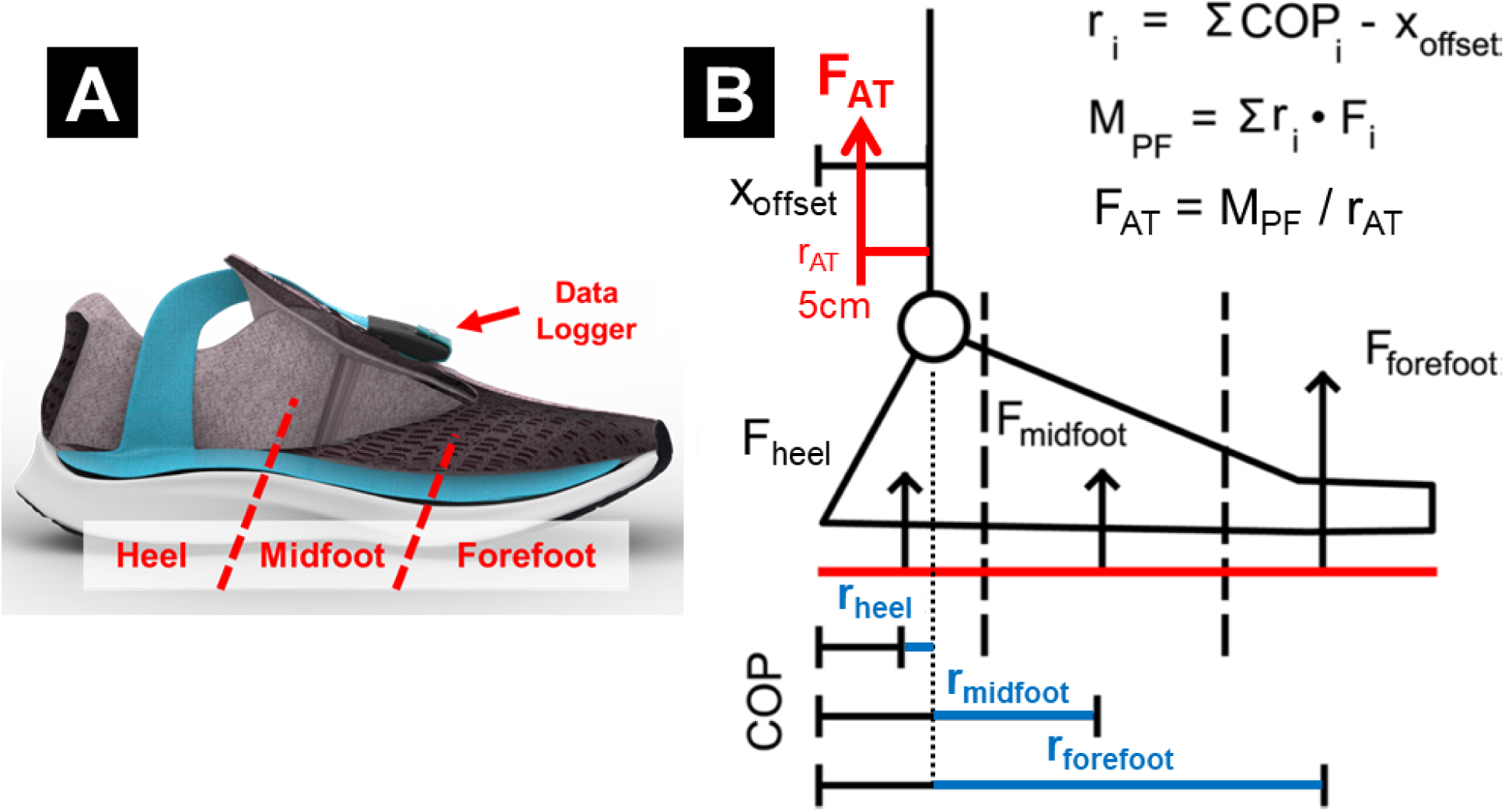
Instrumented insole for Achilles tendon load monitoring. **(A)** Insole device embedded with 3 plantar force sensors, shown here fit into a typical athletic shoe. (This diagram is created with copyright permission from Novel GmbH.) **(B)** Our validated algorithm to estimate Achilles tendon load from the 3 sensor forces. We assumed each force to be perpendicular to the sole and applied at constant center of pressure (COP). We measured the moment arms of each force (r_i_) relative to the ankle joint, computed plantarflexion moment (M_PF_) by the sum of 3 force-by-moment arm products, then estimated Achilles tendon load (F_AT_) by dividing the moment by an assumed tendon moment arm (r_AT_ = 5 cm).

We fit one insole into each participant’s own shoe under the (more) symptomatic foot. We trained participants to initialize the insole each morning, by setting the sensor force to zero when standing on the opposite foot, with a provided controller smartphone for data collection over the following two weeks. We asked participants to wear the insole as much as possible throughout the day and recharge the sensors overnight with a provided cable. We also asked participants to move the insole into any shoe they change into (unless it is infeasible) and re-initialize immediately after shoe change. This means there may be one or multiple data recordings per day of insole wearing, depending on the shoe changes. Over the two-week span, we sent daily text messages to their smartphone reminding them of their insole monitoring experiment. We provided a prepaid return mailing box, which the participants used to mail the instrumented insole back to us at the end of the two-week period. All sensor recordings were logged at 20 Hz and transferred from the data logger to our database via the smartphone. A recent study found that sampling insole sensor data at 20 Hz only induces <3% error in peak plantar force compared to 100 Hz, while increasing real-world data capturing capacity by five times [41].

### Cumulative Achilles tendon load estimation

The data quality of each insole recording was numerically and graphically verified before downstream processing. Specifically, we ensured that sensor force stayed zero whenever unloaded by detecting and removing nonphysical baseline signal drifts (**Supplementary Fig. S6**). We also screened for and removed erroneous recordings possibly caused by incorrect sensor initialization or insole misfit in the shoe (**Supplementary Fig. S7 & Supplementary Table**). This verification process was necessary for removing nonphysical artifacts that would cause incorrect estimation of the Achilles tendon load. Notably, we had to exclude 3 of our 19 initially recruited participants from data analysis because most of their recordings were erroneous. We additionally excluded 1 participant who did not follow our full-day insole monitoring protocol (and instead recorded only 1–3 hours each day), leaving data from 15 participants remaining for analysis.

Achilles tendon load was estimated from sensor-measured plantar contact forces using our validated physics-based algorithm [20]. Briefly, we computed the net moment of plantar force using the 3 insole sensor forces and the size-specific dimensions of each sensor relative to the ankle joint (**Fig. 5B**). This plantarflexion moment was converted into an estimated Achilles tendon load by assuming a 5cm tendon moment arm [42]. Our published study [20] showed that this simple algorithm estimates ankle moment with strong agreement (>96.4%) against standard in-lab motion capture. We normalized this estimated Achilles tendon load by body weight (unit: ×BW) to compare across study participants.

We developed two new measures to quantify the cumulative effect of insole-based Achilles tendon load: the *overall load* and the *high-level load* (**Fig. 6**). First, we summed the cumulative impulse of tendon load through the entire monitoring duration using numerical integration (unit: ×BW×hour), which represents the combined effect of loading magnitude and frequency. Next, we selected two loading magnitude thresholds to delineate a general *overall* level and a representative *high level* of tendon loading, according to our published database on Achilles tendon load during exercises [7]. Specifically, we defined 0.3 times bodyweight (0.3×BW) as the *overall* tendon loading threshold (**Fig. 6A**), because this is the load experienced during the lowest-loading Achilles tendon exercise: seated heel raises [7]. The 0.3×BW threshold thus represents a clinically meaningful baseline tendon load. We defined time above 0.3×BW as each participant’s total loading time with the insole, which excluded all periods of inactivity, insole non-wear, and sensor noises below the baseline (**Supplementary Fig. S8**). Similarly, we defined 3 times bodyweight (3.0×BW) as the *high-level* tendon loading threshold (**Fig. 6B**), because this is approximately the peak Achilles tendon load during walking [7,43]. The 3.0×BW threshold represents a boundary beyond daily living mobility tasks – which is primarily walking – such that cumulative loading above this level results predominantly from dynamic exercises and thus is uniquely relevant to physically active individuals.

**Figure 6.**
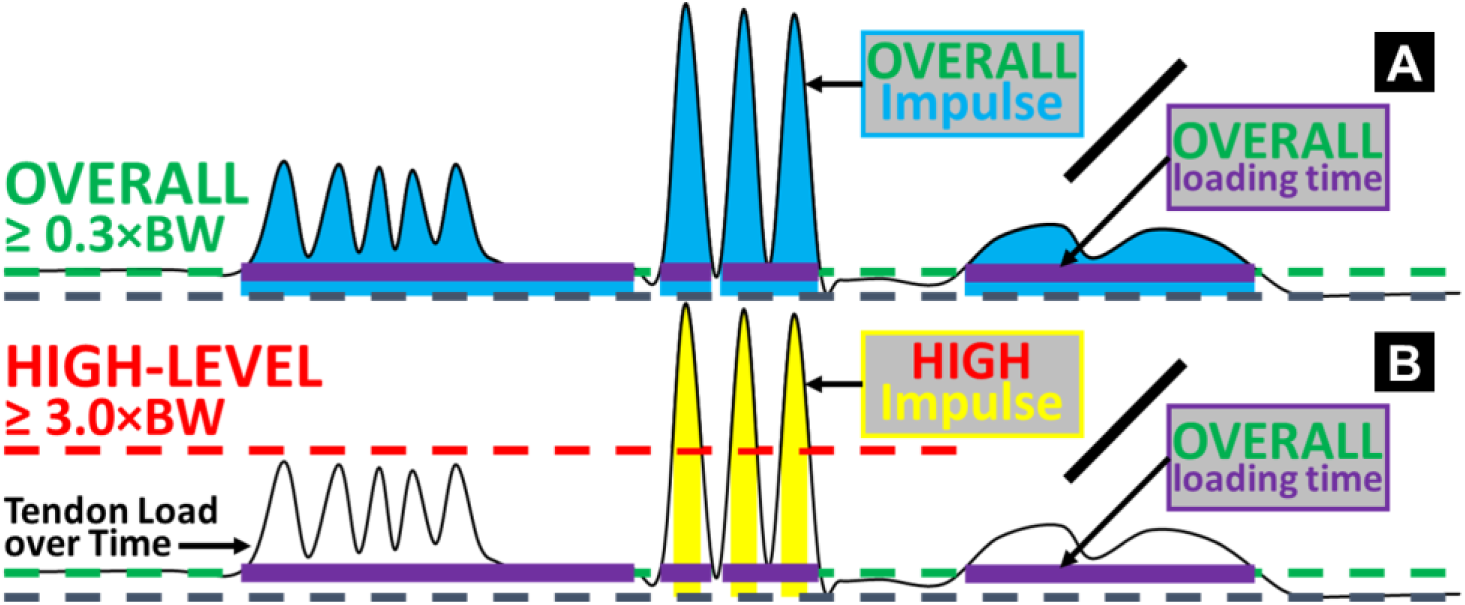
Diagrams illustrating the definitions of the two cumulative Achilles tendon loads, above overall and high-level thresholds. The waveform in each diagram exemplifies insole-based Achilles tendon load (vertical axis) as a function of time (horizontal axis). **(A)** We defined an overall threshold of 0.3×BW to represent loading incurred from any daily living activity that loads the Achilles tendon. This excluded inactivity periods and whenever the sensor was recording but not worn (e.g., right end of the diagram). We defined overall cumulative time above 0.3×BW (purple bars) as the total loading time with the insole. We summed loading impulse above this overall level (blue areas) and divided it by the total loading time for the overall tendon load accumulated *per hour*. **(B)** Similarly, we defined a high-level threshold of 3.0×BW to quantify loading incurred beyond walking and thus primarily resulted from dynamic physical activities. We summed loading impulse above this high-level threshold (yellow areas) and divided it by the total loading time (*not high-level* cumulative time) for the high-level tendon load accumulated *per hour*.

These two measures of the cumulative *overall* and *high-level* tendon load were not readily proportional to the full amounts of real-world loading, because they were still dependent on insole wear time. Namely, the total time a study participant is able or willing to wear the insole does not exactly correspond to the time when that individual is physically active and loading their Achilles tendon. Practical considerations like the type of shoes worn, preference about at-home wearing, bedtime variability, and concerns about aesthetics are some reasons participants may have chosen not to wear the insole during parts of the two-week period. To account for these sources of variability, we divided the cumulative overall and high-level loads by the total time spent above the overall load level, i.e., participant’s total loading time with the insole (**Fig. 6**). This yielded the *normalized* overall and high-level loads accumulated *per hour* (unit: ×BW). We used these two *normalized* measures of cumulative Achilles tendon loading on each of the 15 participants for statistical analysis.

### Statistical analysis

We computed Pearson correlations between insole-based cumulative Achilles tendon loads (overall and high-level) with dynamometer-based plantar flexor functional capacity, motion capture-based dynamic function, and survey-based clinical outcome measures (**Table 2**; n = 15 for each correlation test). We tested two measures of insole-based cumulative Achilles tendon load:

**Table 2.** Measurement categories for statistical analysis (*technique used*), the specific measures tested, and their units.

|  |  |  |
| --- | --- | --- |
| **Category** | **Measures** | **Unit** |
| **Achilles tendon cumulative loading**<br>(*Insole sensors*) | (1) Overall tendon load ($\geq 0.3 \times BW$)<br>*normalized per hour* | $\times BW$ |
| | (2) High-level tendon load ($\geq 3.0 \times BW$)<br>*normalized per hour* | |
| **Plantar flexor functional capacity**<br>(*Dynamometer testing*) | (1–3) Peak plantar flexor moment<br>(isometric, $30^\circ/s$, $150^\circ/s$) | $\% H \times W$ |
| | (4–5) Peak plantar flexor power<br>($30^\circ/s$, $150^\circ/s$) | $\% H \times W \times rad/s$ |
| | (6–7) Mechanical work per contraction<br>($30^\circ/s$, $150^\circ/s$) | $\% H \times W \times rad$ |
| **Plantar flexor dynamic function**<br>(*Motion capture*) | (1) Double-leg heel raise height<br>(more symptomatic side) | $\% BH$ |
|  | (2) Single-leg countermovement jump height |  |
| | (3) Single-leg heel raise repetitions on $25^\circ$ slope | – |
| **Self-reported outcomes and characteristics**<br>(*Survey questionnaires*) | (1) Age | Years |
|  | (2) VISA-A score (more symptomatic side) | – |
|  | (3) PAS score (current level) | – |
| ×, times; BW/W, Body Weight; BH/H, Body Height; $^\circ$, degree; s, second; rad, radius; VISA-A, Victorian Institute of Sports Assessment – Achilles questionnaire; PAS, Physical Activity Scale. | | |

(1) normalized overall load *per hour* and (2) normalized high-level load *per hour*. We tested seven measures of plantar flexor functional capacity: (1–3) peak moment during isometric, slow (30°/s) isokinetic, fast (150°/s) isokinetic contractions, (4–5) peak power during slow and fast isokinetic contractions, (6–7) mechanical work per slow and fast isokinetic contraction. We tested three measures of plantar flexor dynamic function: (1) double-leg heel raise height on the (more) symptomatic side, (2) single-leg countermovement jump height, (3) repetitions of single-leg heel raise on the inclined slope. We tested three survey-based measures: (1) age, (2) VISA-A score on the (more) symptomatic side, (3) current Physical Activity Scale. We defined the strengths of Pearson correlation according to literature [23,24]: |r| ≥ 0.6 as strong, 0.4 – 0.6 as moderate, and < 0.4 as weak. We additionally defined the significance level of all correlations at *α* = 0.05. Statistical significance of the Pearson correlations was used only as an ancillary metric, because we consider the strengths of correlations (weak, moderate, or strong) as our primary statistical results.

### Insole data subsampling analysis

We performed an ancillary analysis to determine the minimal amount of insole data needed to reliably represent habitual two-week Achilles tendon loading. This analysis is important due to the practical challenges regarding using insole sensors, and useful for establishing best practices that future studies can leverage to balance data fidelity with participant burden. We iteratively quantified the difference between our full two-week insole dataset and extracted data subsamples with a reduced number of monitoring days. Specifically, using *overall* and *high-level* cumulative tendon loads estimated from the full two-week dataset as reference, we computed the mean absolute percent error (percent difference from full dataset-based) [18,44,45] and Pearson correlations when estimating from gradually reduced days of insole data: 6 (the first six days recorded by each participant), 5 (omit day-1,…, omit day-6), 4, 3, 2, or 1 day (day-1 only,… day-6 only). We hypothesized that cumulative tendon loads based on 6-day insole data would remain close to full dataset-based estimates, whereas using 1 day of data would lead to larger differences.

## Data Availability

The insole sensor data, dynamometer testing data, motion capture data, deidentified survey data, and data analysis code that we generated and/or analyzed during the current study are openly and freely available in a public Zenodo repository [46], https://doi.org/10.5281/zenodo.14947735, which we shared in accordance with study participant protection standards.

## Data Availability

The insole sensor data, dynamometer testing data, motion capture data, deidentified survey data, and data analysis code that we generated and/or analyzed during the current study are openly and freely available in a public Zenodo repository, https://doi.org/10.5281/zenodo.14947735, which we shared in accordance with study participant protection standards.

## Acknowledgements / Funding

Research reported in this study was supported by the National Institute of Arthritis and Musculoskeletal and Skin Diseases of the National Institutes of Health, under grant numbers R01AR078898 and P50AR080581. The funder played no role in study design, data collection, analysis and interpretation of data, or the writing of this manuscript. The content is solely the responsibility of the authors and does not necessarily represent the official views of the National Institutes of Health. The authors thank Devyn P. Russo for assistance with data collection.

## Author Contributions

K.S., K.G.S. and J.R.B. conceptualized the study. J.R.B. acquired funding and resources for the study and supervised project administration. K.S., R.T.P., K.G.S. and J.R.B. developed the methodology. K.S., M.P.K. and A.K.S. curated experimental data. K.S. and M.P.K. performed hardware and software validation and formal analysis of the data. K.S. developed the original draft of the manuscript, including visualization of the analyzed data. All authors reviewed and edited the manuscript and approved the final version submitted to the journal.

## Supplementary Information

The online version of this article contains supplementary material available.

