## Supplementary Information for "Two weeks of cumulative tendon load monitored by insole sensors is associated with plantar flexor function in Achilles tendinopathy"

**Supplementary Figure S1.** *Plantar flexor functional capacity: dynamometer data*

Plantar flexor functional capacity varied among the study participants and depended upon the dynamometer testing condition. As a group, participants produced peak plantarflexion moment of 2.4%H×W (Standard Deviation, SD: 1.1%H×W) during isometric contraction, 2.4%H×W (SD: 1.0%H×W) during slow isokinetic contraction (30°/s), and 1.0%H×W (SD: 0.6%H×W) during fast isokinetic contraction (150°/s) (**Supplementary Fig. S1A**). Participants produced peak plantarflexion power 1.3%H×W×rad/s during slow isokinetic contraction (SD: 0.6%H×W×rad/s) and 2.5%H×W×rad/s during fast isokinetic contraction (SD: 1.4%H×W×rad/s) (**Supplementary Fig. S1B**). Per contraction, participants produced mechanical work of 1.1%H×W×rad during slow isokinetic (SD: 0.6%H×W×rad) and 0.6%H×W×rad during fast isokinetic contraction (SD: 0.4%H×W×rad) (**Supplementary Fig. S1C**). In general, fast isokinetic contraction produced smaller peak plantarflexion moment and work per contraction compared to isometric or slow isokinetic contraction, but larger peak plantarflexion power due to its higher speed of motion.

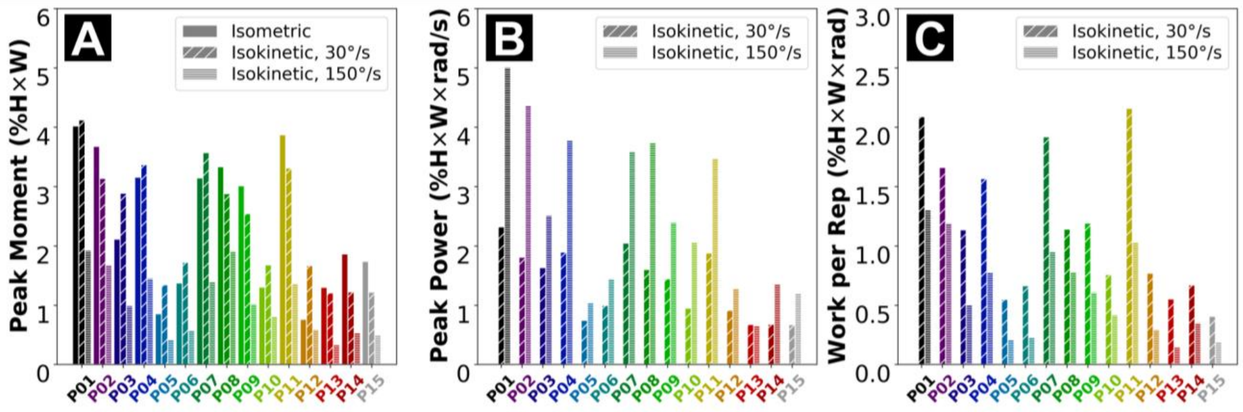


**Supplementary Figure S1**. Plantar flexor function quantified in peak moment **(A)**, peak power **(B)**, and total mechanical work per repetition **(C)** during the 3 testing conditions for each of our 15 study participants (P##, sorted in the order of ascending age). Each metric was normalized by the participant's body height time weight and averaged over 3 successful repetitions. For isometric contraction, only the peak moment was reported because no mechanical power or work was produced during this static testing condition.

**Supplementary Figure S2.** *Subsampling: non-normalized errors and scatter graphs*

Reduced days of insole data led to increased error in the cumulative overall and high-level tendon loads, with decreased correlations, to the full dataset-based estimate. Non-normalized mean absolute errors (in ×BW unit) versus days of data used are reported in **Supplementary Fig. S2** below. Mean absolute error and correlation coefficient for each single day and each specific multi-day combination (1–6 days) are reported throughout **Supplementary Fig. S2.1–S2.6**.

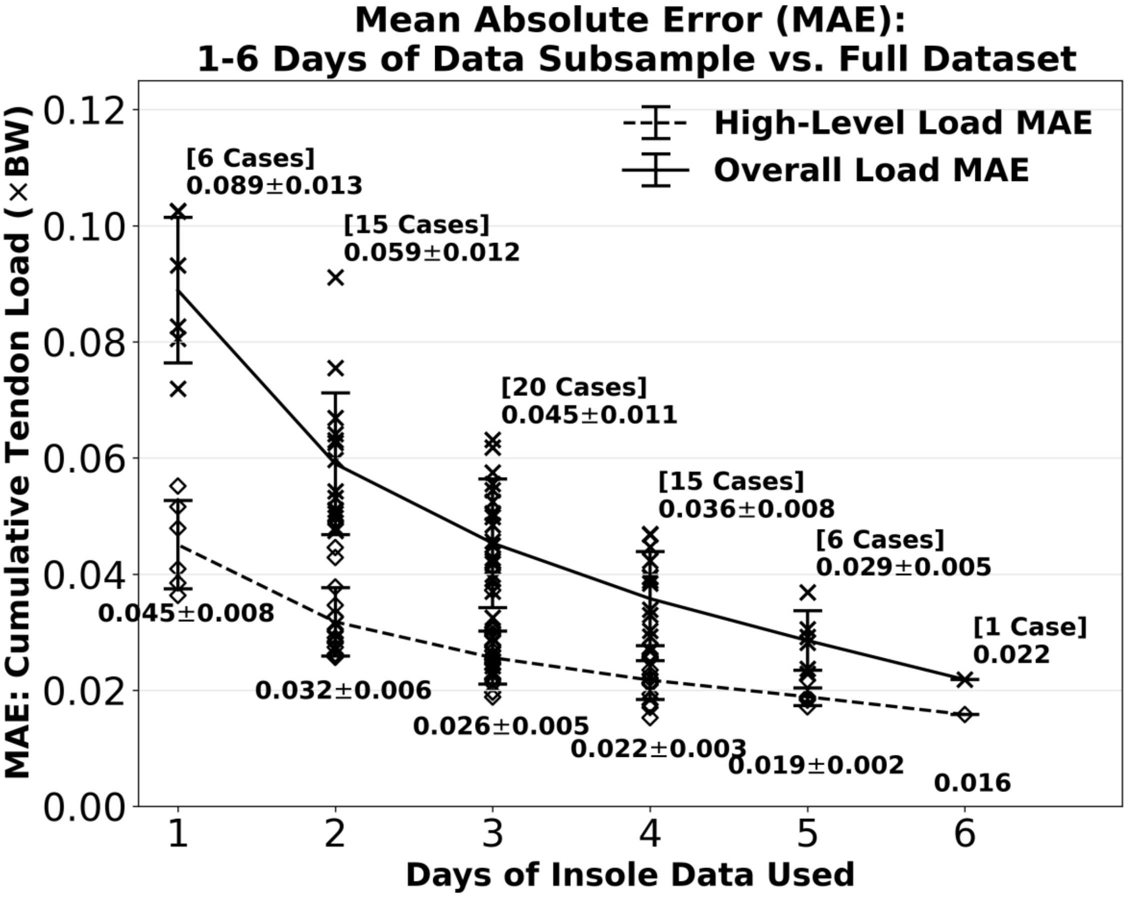


**Supplementary Figure S2.** Mean absolute error (unit: ×BW) for cumulative overall and high-level tendon loads estimated from insole data subsamples. Error for 1 day-based estimate was 0.089×BW from full dataset-based for the overall load, and 0.045×BW for the high-level load. When using 6 days of data, mean absolute error for the overall load was reduced to 0.022×BW and the high-level load to 0.016×BW. Error bars represent ±1 standard deviation from the mean across all 1, 2, …, or 6 day-based estimates.

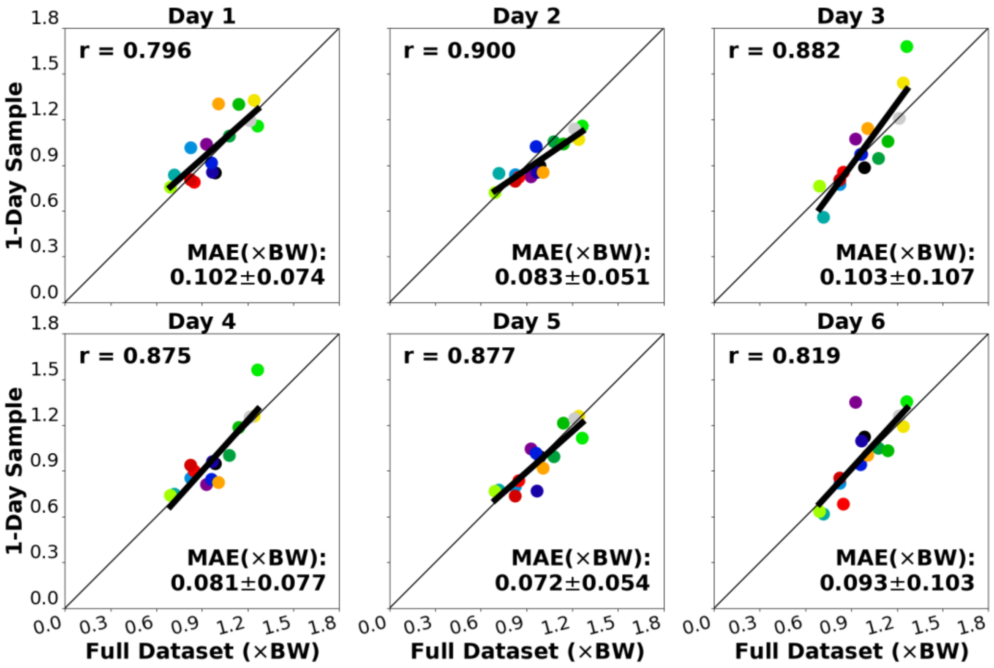


**Supplementary Figure S2.1.A.** 1-day estimate of overall cumulative Achilles tendon load (vertical axis) correlated to the full two-week dataset-based estimate (diagonal). Each dot represents a participant.

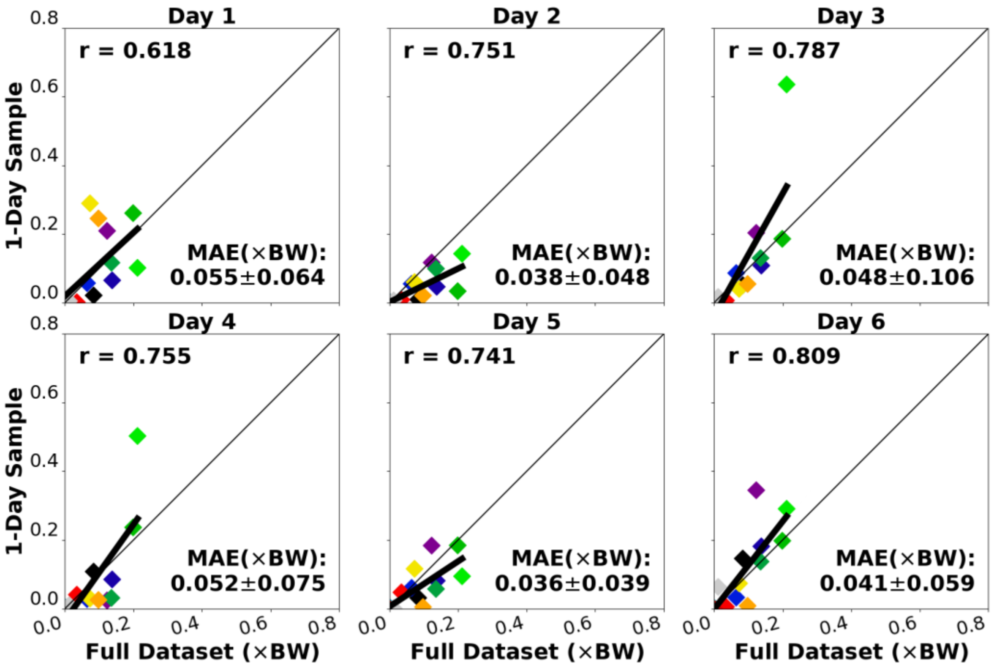


**Supplementary Figure S2.1.B.** 1-day estimate of high-level cumulative Achilles tendon load (vertical axis) correlated to the full two-week dataset-based estimate (diagonal). Each dot represents a participant.

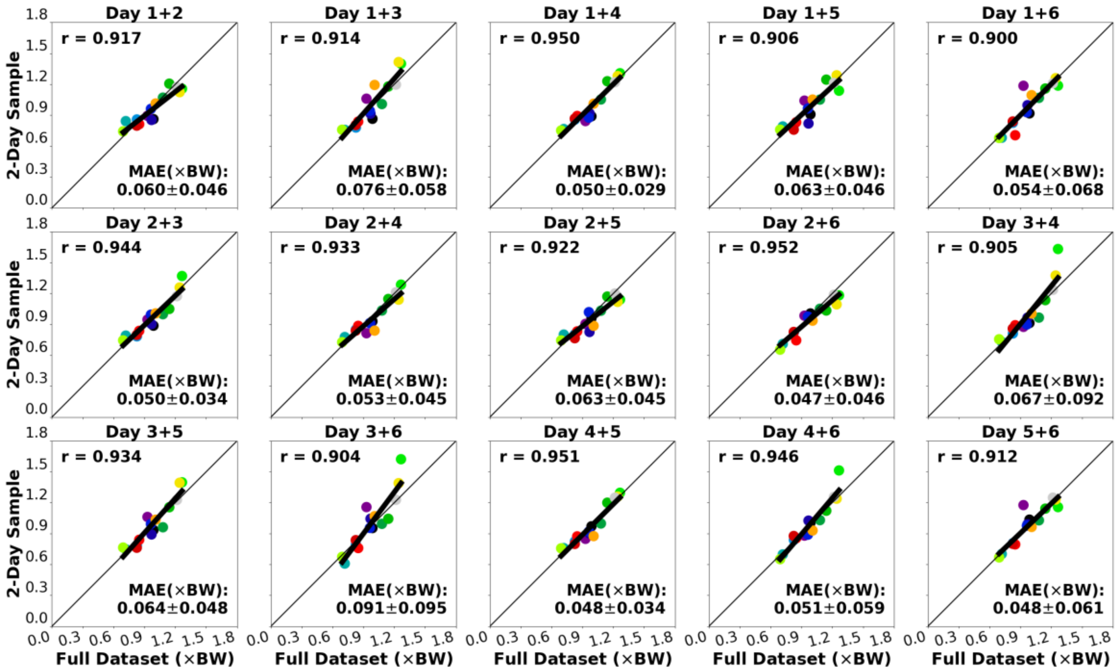


**Supplementary Figure S2.2.A.** <u>2-day</u> estimate of <u>overall</u> cumulative Achilles tendon load (vertical axis) correlated to the full two-week dataset-based estimate (diagonal). Each dot represents a participant.

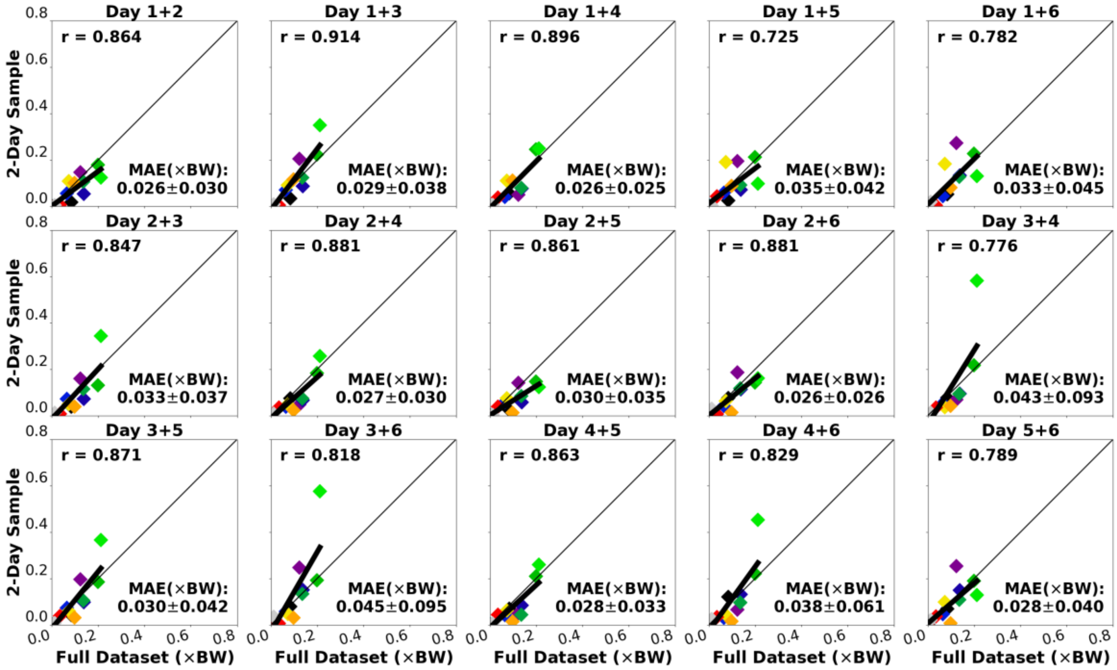


**Supplementary Figure S2.2.B.** <u>2-day</u> estimate of <u>high-level</u> cumulative Achilles tendon load (vertical axis) correlated to the full two-week dataset-based estimate (diagonal). Each dot represents a participant.

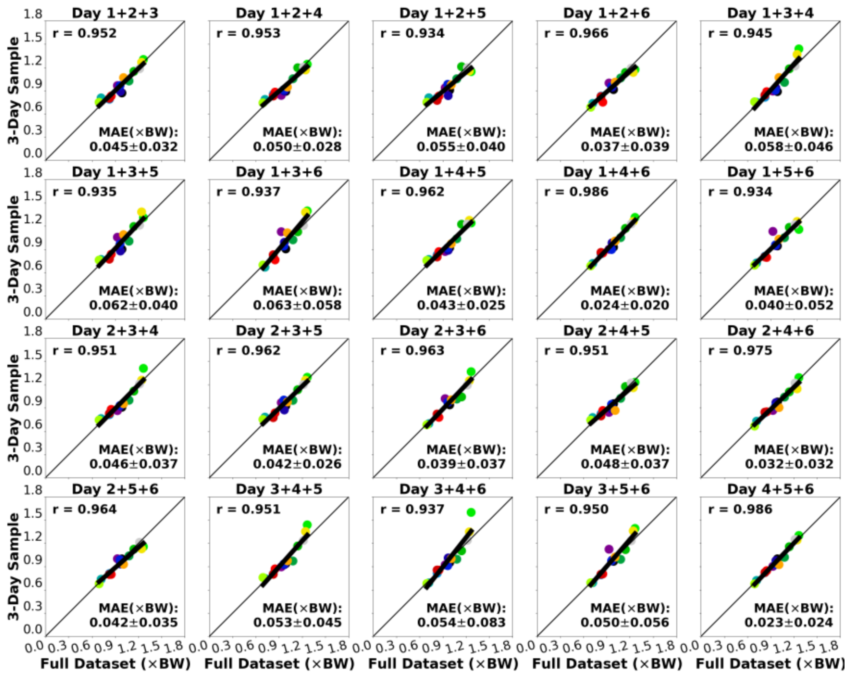


**Supplementary Figure S2.3.A.** 3-day estimate of overall cumulative Achilles tendon load (vertical axis) correlated to the full two-week dataset-based estimate (diagonal). Each dot represents a participant.

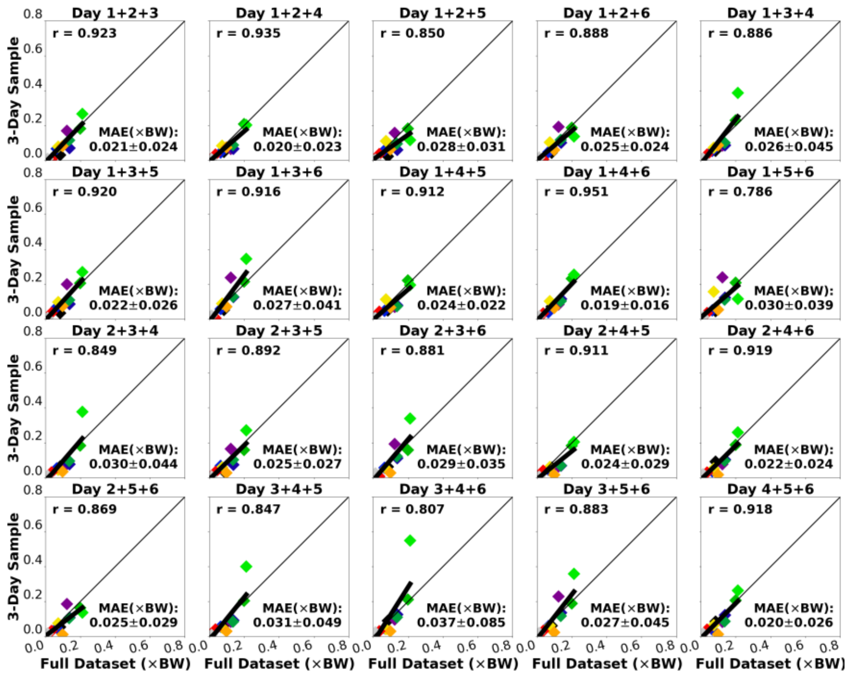


**Supplementary Figure S2.3.B.** 3-day estimate of high-level cumulative Achilles tendon load (vertical axis) correlated to the full two-week dataset-based estimate (diagonal). Each dot represents a participant.

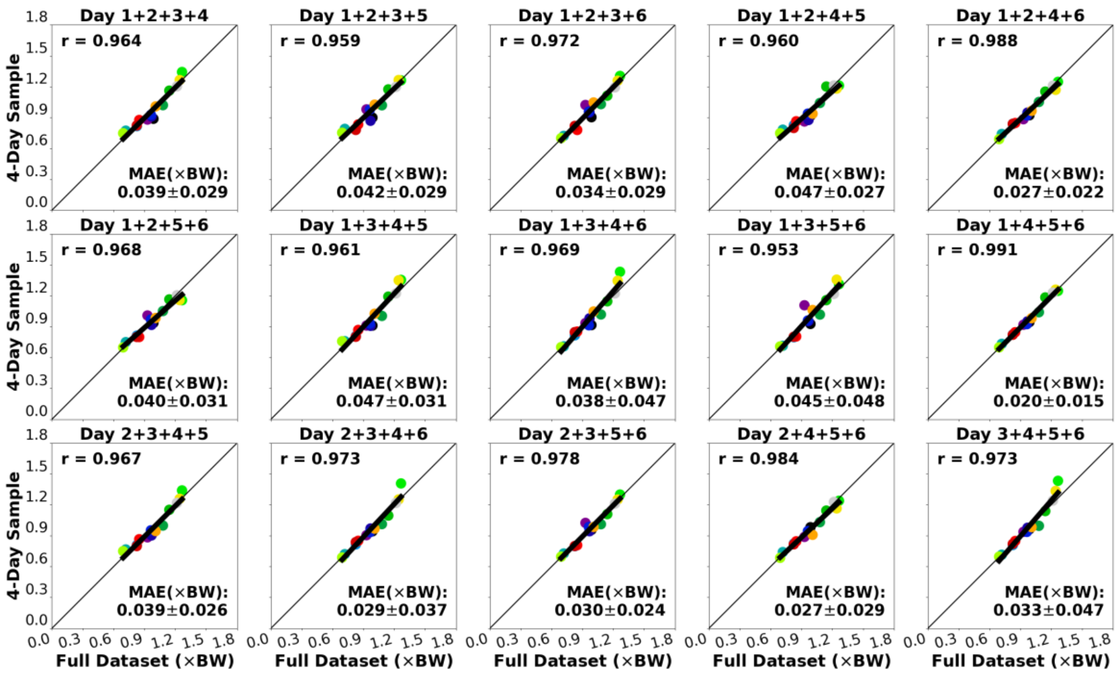


**Supplementary Figure S2.4.A.** 4-day estimate of overall cumulative Achilles tendon load (vertical axis) correlated to the full two-week dataset-based estimate (diagonal). Each dot represents a participant.

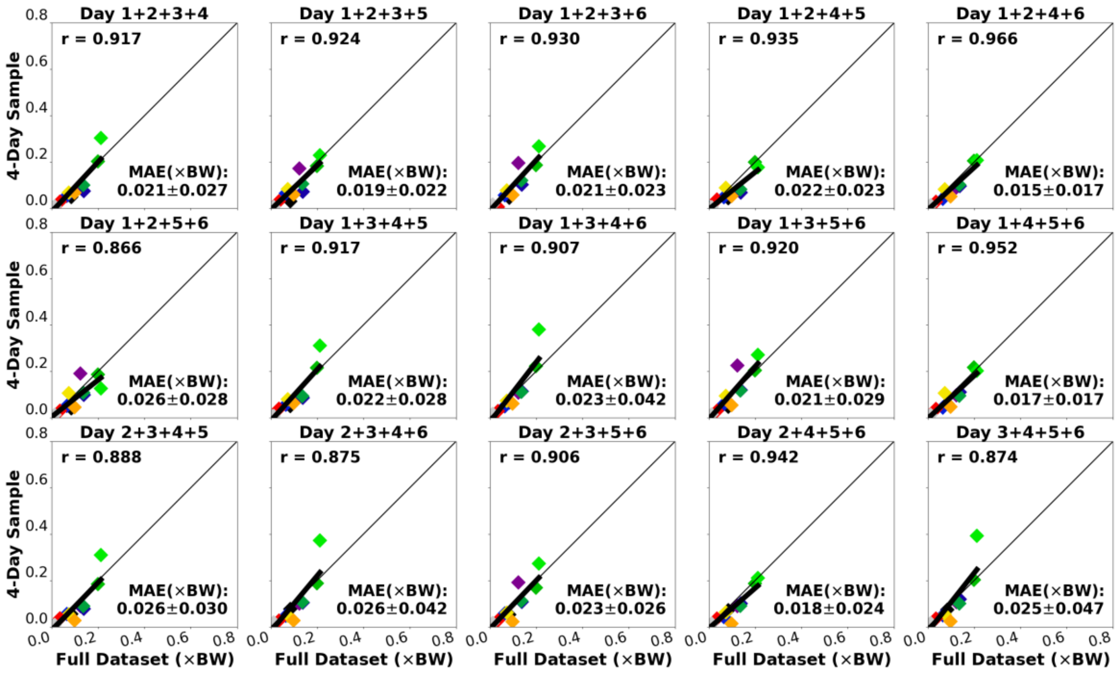


**Supplementary Figure S2.4.B.** 4-day estimate of high-level cumulative Achilles tendon load (vertical axis) correlated to the full two-week dataset-based estimate (diagonal). Each dot represents a participant.

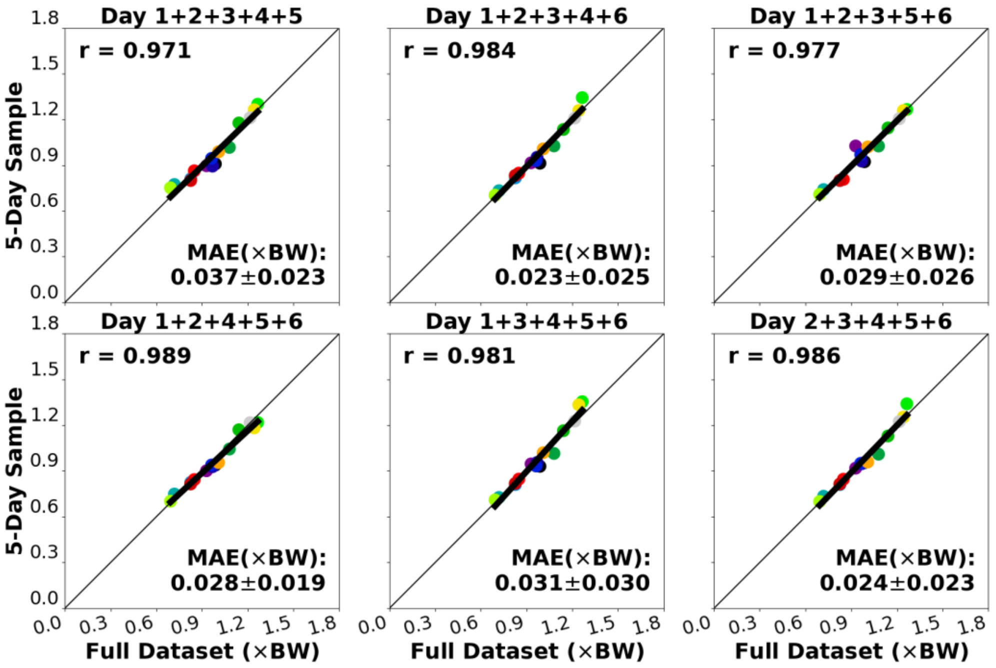


**Supplementary Figure S2.5.A.** <u>5-day</u> estimate of <u>overall</u> cumulative Achilles tendon load (vertical axis) correlated to the full two-week dataset-based estimate (diagonal). Each dot represents a participant.

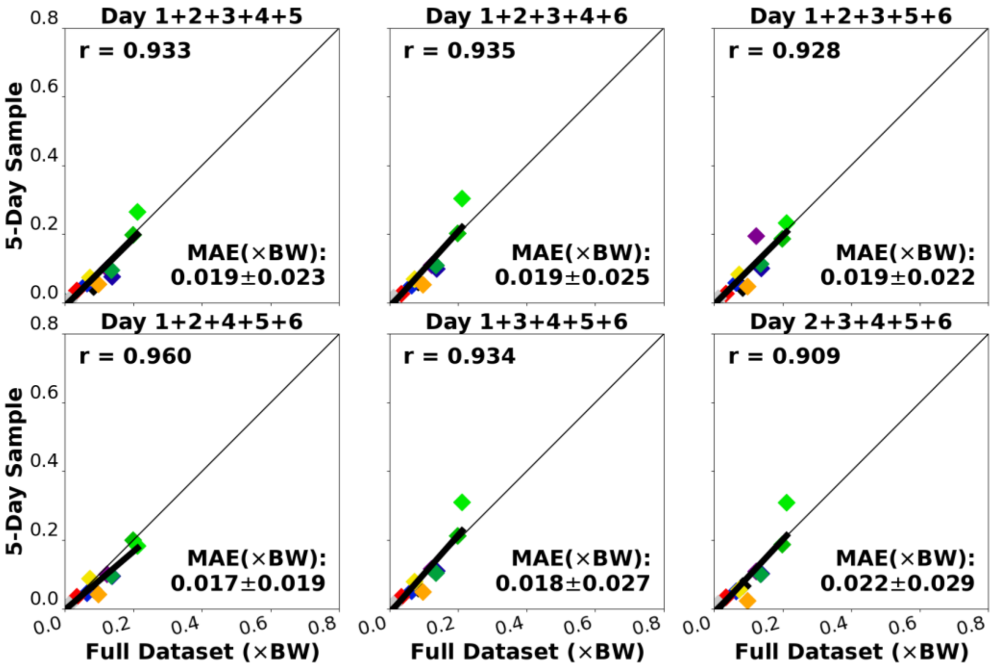


**Supplementary Figure S2.5.B.** <u>5-day</u> estimate of <u>high-level</u> cumulative Achilles tendon load (vertical axis) correlated to the full two-week dataset-based estimate (diagonal). Each dot represents a participant.

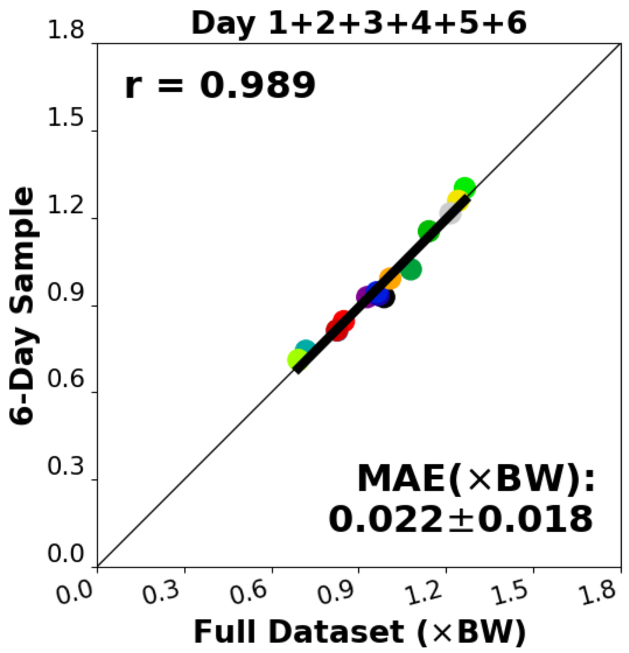


**Supplementary Figure S2.6.A.** 6-day estimate of overall cumulative Achilles tendon load (vertical axis) correlated to the full two-week dataset-based estimate (diagonal). Each dot represents a participant.

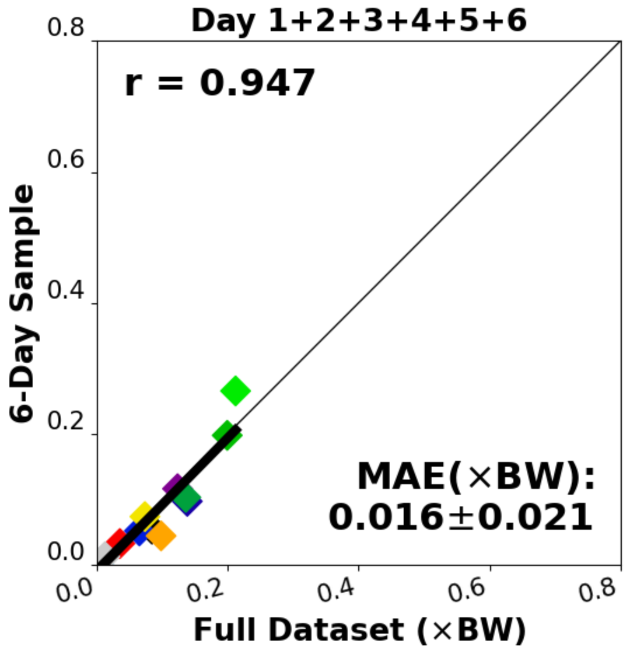


**Supplementary Figure S2.6.B.** 6-day estimate of high-level cumulative Achilles tendon load (vertical axis) correlated to the full two-week dataset-based estimate (diagonal). Each dot represents a participant.

**Supplementary Figure S3.** *Dynamometer (functional capacity) vs. survey measures*

Most dynamometer-based plantar flexor functional capacity measures were moderate-to-strongly and negatively correlated to age, as we expected (r = -0.370 – -0.640; **Supplementary Fig. S3A**). Yet, all dynamometer-based plantar flexor function metrics had only weak positive correlations with the VISA-A score (r = 0.217–0.301; **Supplementary Fig. S3B**). Likewise, dynamometer-based plantar flexor function metrics were only weak-to-moderately albeit positively correlated to self-reported activity level (r = 0.329–0.491; **Supplementary Fig. S3C**).

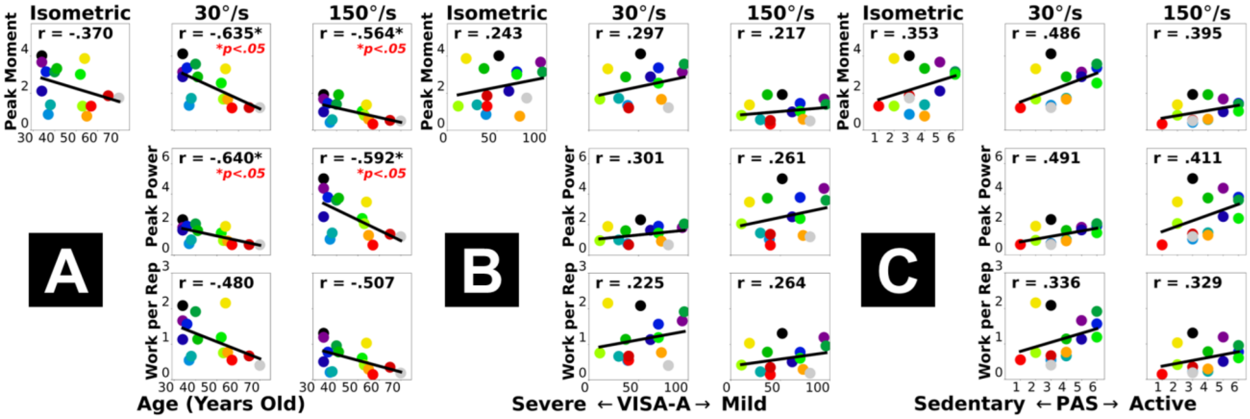


**Supplementary Figure S3.** Pearson correlations between dynamometer-based plantar flexor functional capacity (vertical axis) and survey-based measures of study participant characteristics or outcomes (horizontal axis): age **(A)**, severity of Achilles tendinopathy – VISA-A score **(B)**, and self-reported current activity level – PAS score **(C)**. Each dot represents a participant. "*" annotates statistical significance of the correlation ($p < 0.05$).

**Supplementary Figure S4.** *Motion capture (dynamic function) vs. survey measures*

Motion capture-based plantar flexor dynamic function measures were negatively correlated to age and increasingly with the dynamic demand of the task (**Supplementary Fig. S4A**), with a strong correlation for the challenging single-leg heel raises on the 25° slope (r = -0.760). Yet, consistent with dynamometer-based measures, the plantar flexor dynamic function also had weak-to-moderately positive correlations with the VISA-A score (r = 0.080–0.419; **Supplementary Fig. S4B**) and the self-reported activity level (r = 0.227–0.491; **Supplementary Fig. S4C**).

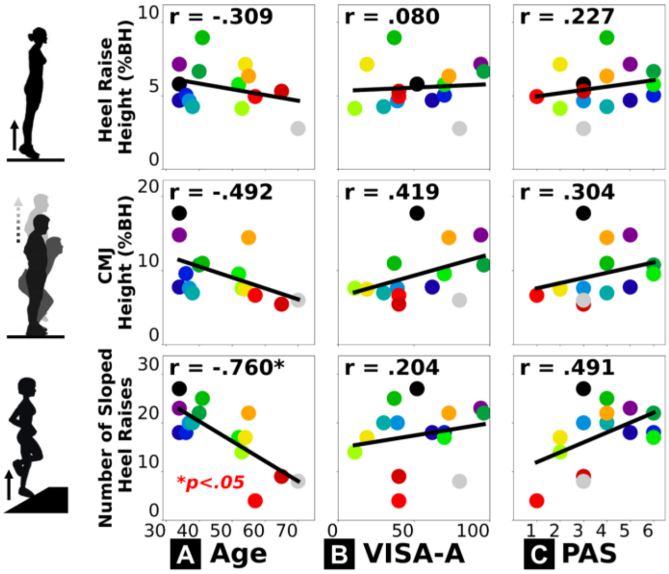


**Supplementary Figure S4.** Pearson correlations between motion capture-based plantar flexor dynamic function (vertical axis) and survey-based measures of study participant characteristics or outcomes (horizontal axis): age **(A)**, severity of Achilles tendinopathy – VISA-A score **(B)**, and self-reported current activity level – PAS score **(C)**. Each dot represents a participant. "*" annotates statistical significance of the correlation ($p < 0.05$).

**Supplementary Figure S5.** *Plantar flexor function: dynamometer vs. motion capture*

Plantar flexor functional capacity assessed with dynamometer were moderately-to-strongly correlated to dynamic function assessed with motion capture, across all 3 dynamic movements we tested (r = 0.462 – 0.703; **Supplementary Fig. S5**). All correlations were significant for the 2 isokinetic testing conditions (middle and right columns in each panel), plus isometric strength with the double-leg heel raise height (**Supplementary Fig. S5A**, top left). As we expected, functional capacity of the plantar flexors during strength testing is closely related to their dynamic functional performance during exercises.

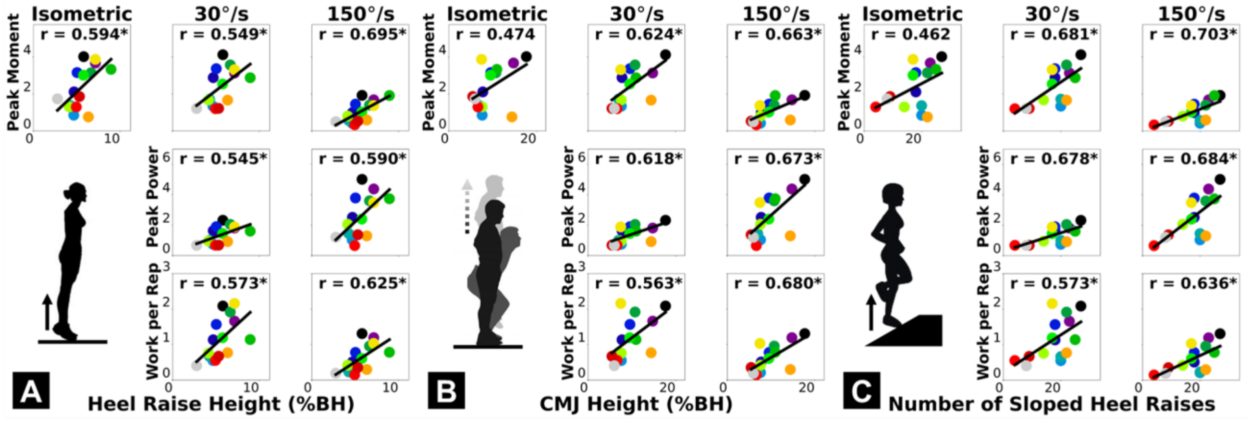


**Supplementary Figure S5.** Pearson correlations between dynamometer-based plantar flexor functional capacity (vertical axis) and motion capture-based plantar flexor dynamic function (horizontal axis): double-leg heel raise height on the (more) symptomatic side **(A)**, single-leg countermovement jump height **(B)**, and repetitions of single-leg heel raise on the inclined slope **(C)**. Each dot represents a participant. Pearson correlations were statistically significant for all isokinetic measures of functional capacity (30°/s and 150°/s; "*" annotates $p < 0.05$).

**Supplementary Figure S6.** *Detecting and removing insole sensor signal drift*

Participants initialized insole sensors each morning by setting the plantar forces to zero when standing on the other foot to unload the insole. However, for the insole sensors we used, this zero level sometimes became inaccurate over the day and caused the force baseline to drift off zero (**Supplementary Fig. S6**). To remove this nonphysical drift, we built a bi-directional filter that detects the 10-minute rolling minimum of the force, which we assumed to be the true zero and subtracted from raw data for each sensor. We chose this 10-minute window as the shortest realistic duration in which a true zero-force event surely exists: the foot should lift off ground at least once. We computed Achilles tendon load using the 3 sensor signals filtered through this "drift remover".

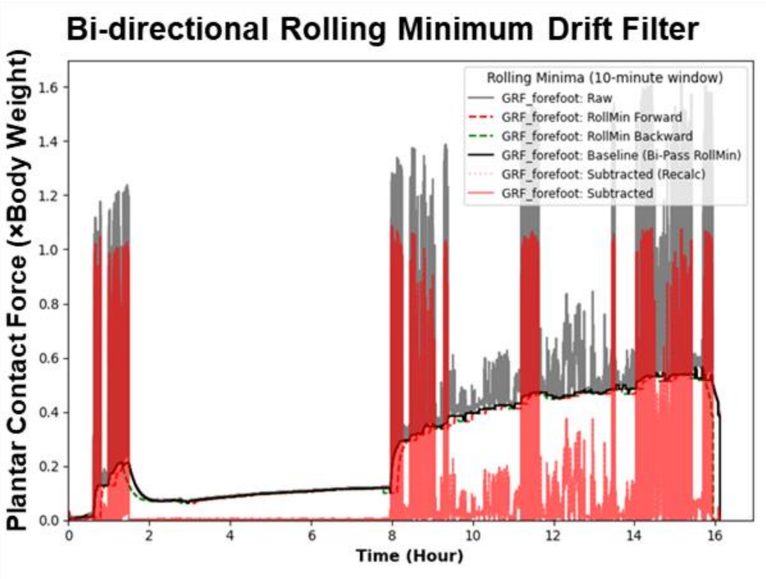


**Supplementary Figure S6**. Bi-directional rolling minimum filter to remove insole sensor baseline drift. In our sensors, the original sensor force (gray) tended to drift off zero overtime. By detecting and subtracting the bi-directional rolling minimum (both forward and backward) over a 10-minute window, we were able to find and subtract this nonphysical baseline (black) and extract the actual plantar forces over the day (red).

**Supplementary Figure S7 & Supplementary Table.** *Summary of insole recording errors*

Although we were able to correct for baseline drifts (**Supplementary Fig. S6**), two other types of error found in unprocessed insole recordings were uncorrectable and we had to remove the affected data from further processing. (1) Incorrect scaling factor (electric signal-to-force ratio) that distorted the measured force by an unknown nonphysical factor (**Supplementary Fig. S7A**). (2) Unrealistically high force magnitude (e.g., in orders of $10^4$–$10^5$ N and >100×BW), possibly due to insole misfit in the shoe short-circuiting sensor wires (**Supplementary Fig. S7B**). The number of unprocessed "raw" insole recordings, corrupted files by error type, and final number of "good" recordings used for analysis are summarized below (**Supplementary Table**).

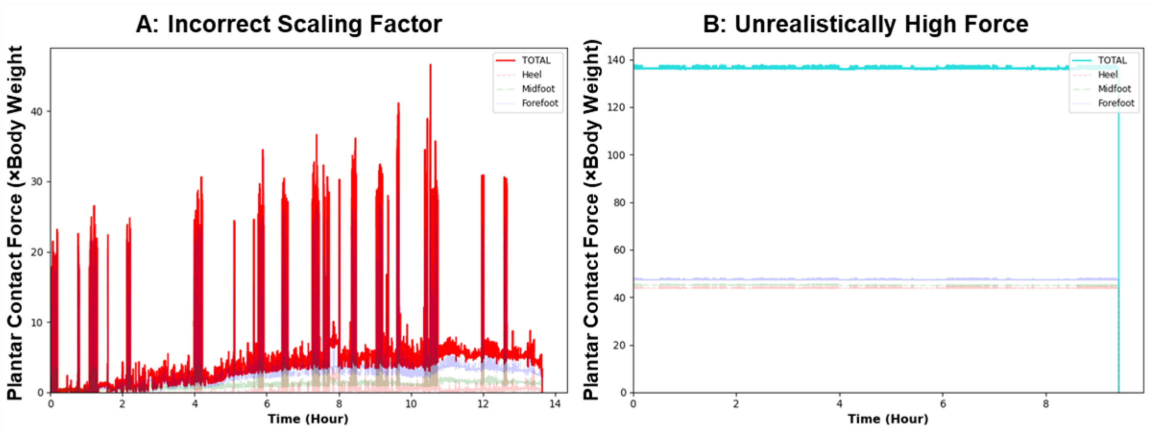


**Supplementary Figure S7**. Example of two types of uncorrectable insole recording error that warranted data removal: **(A)** incorrect scaling factor (~40×BW) and **(B)** unrealistically high force (>100×BW).

| **Supplementary Table.** Number of unprocessed, removed, and used insole recordings. File counts below combine all 15 included participants, who may record none, one, or multiple data files each day. | |
|---|---|
| Total number of unprocessed "raw" insole recording files | 175 Files |
| Unrealistically high force: uncorrectable, removed (**Supplementary Fig. S7B**) | 3 Files |
| Incorrect scaling factors: uncorrectable, removed (**Supplementary Fig. S7A**) | 2 Files |
| **Number of "good" recordings used for downstream data analysis** | **170 Files** |
| *Nonphysical baseline drift: corrected (**Supplementary Fig. S6**)* | *37 Files* |
| *Number of true error-free insole recording files* | *133 Files* |

**Supplementary Figure S8.** *Threshold to exclude sensor noises, non-wear, and inactivity*

We computed cumulative Achilles tendon load as the integration of load over time. Yet the highly variable durations of recording, including when the insole was not worn, caused cumulative tendon load to have no clear associations with the recording time (**Supplementary Fig. S8A**). By using the 0.3×BW overall tendon load as the cutoff threshold for insole wear, we excluded baseline insole sensor noises and all periods of sensor non-wear and inactivity (e.g., seated). Time spent above this loading threshold (i.e., tendon-loading time with the insole) is much better associated with cumulative Achilles tendon load (**Supplementary Fig. S8B**).

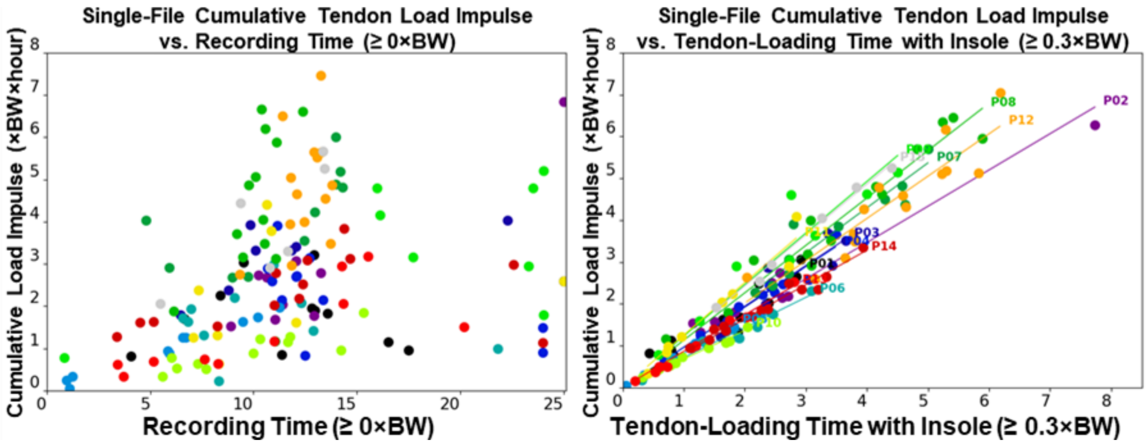


**Supplementary Figure S8**. Single-file cumulative Achilles tendon load impulse (vertical axis, *non-normalized*, unit: ×BW×hour) versus **(A)** recording time (above 0×BW) and **(B)** tendon-loading time with the insole (above 0.3×BW). Each color represents a participant (P##), while each dot represents data from one insole recording file. **(A)** Highly variable recording durations caused cumulative tendon load to have no clear associations with the recording time. **(B)** Tendon-loading time above the 0.3×BW cutoff threshold shows much better associations with cumulative Achilles tendon load.
